# Factors in measuring lumbar spondylolisthesis with reference data from NHANES-II

**DOI:** 10.1101/2022.08.01.22278280

**Authors:** John A. Hipp, Trevor F. Grieco, Patrick Newman, Vikas V. Patel, Charles A. Reitman

**Affiliations:** Medical Metrics, Imaging Core Laboratory, Houston, TX 77056; Colorado University Orthopedics; Orthopaedics and Physical Medicine, Medical University of South Carolina, Charleston, SC 29425

**Keywords:** Spondylolisthesis, disc height, disc angle, normal, radiographic

## Abstract

**Background:** Although spondylolisthesis, disc height loss, and abnormal disc angles are commonly assessed for in clinical practice and research studies, the factors that influence measurements are incompletely understood, and reference data representing a very large and diverse population are not available. Over 7000 lumbar spine x-rays were collected as part of the 2nd National Health and Nutrition Examination Survey (NHANES-II). This nationwide probability sample may facilitate development of robust reference data to objectively classify spondylolisthesis, disc heights, or disc angles as normal vs abnormal. Analysis of lumbar spine x-rays may also help to document whether disc heights and disc angles must be considered when assessing for spondylolisthesis.

**Methods:** Anatomic landmarks were obtained using neural networks and coded logic for L1 to S1 in the NHANES-II lumbar spine radiographs. Nine disc and sagittal plane offset (SPO) measurements were calculated from the landmarks. These data were trimmed to exclude abnormal discs and SPO. The factors that affect SPO were explored along with associations between the metrics and with age, sex, race, nation of origin and BMI. The prevalence of abnormalities was tabulated. Metrics were also calculated for previously analyzed lumbar flexion-extension x-rays to better understand the dependence of SPO on disc angles and disc heights. The errors that occur due to variability in radiographic projection were also assessed.

**Results:** SPO and disc metrics were obtained for 35,490 levels from 7,358 NHANES-II subjects who were 25 to 74 years old. Descriptive statistics for nine SPO and disc metrics were tabulated before and after trimming the data. Age, BMI, and sex were statistically significant but explained little of the variance in the metrics. SPO was significantly dependent on disc angle and height, though less so in the NHANES radiographs than in lumbar flexion-extension studies. Errors in the metrics due to out-of-plane imaging are generally small but can be large with extensive out-of-plane x-rays.

**Discussion:** The NHANES-II collection of lumbar x-rays allows for establishing reference data for SPO and disc metrics. These reference data allow for easily interpreted standardized reporting in units of std dev from average normal. SPO was significantly dependent on disc angle and disc height although the effect is small when there is limited flexion or extension. If SPO is being assessed from flexion or extension, a simple correction can be made. Caution is needed when interpreting measurements when the x-ray beam passes obliquely through the endplates or posterior wall (out-of-plane imaging).

**Conclusions:** The NHANES-II data may facilitate standardized assessments of SPO, disc heights and disc angles. Adjustments should be applied to SPO measurements if made from x-rays with the patient in flexed or extended positions.

## Introduction

Spondylolisthesis is defined as an <u>abnormal</u> “slip” of one vertebra relative to another and is most commonly assessed in the sagittal plane.^1^ Some amount of “slip” occurs with healthy discs. The challenge is to reliably classify “slip” as normal vs abnormal while accounting for significant co-factors. That requires both a reliable measurement method, reference data defining normal vs abnormal, and an understanding of important cofactors. In clinical practice, spondylolisthesis is often subjectively assessed, due to the time required to carefully make the measurements, and the lack of definitive evidence that quantitative measurements are needed. Definitive evidence for/against quantitative measurements requires yet-to-be-completed research, and the outcome will depend on the quality of the quantitative measurements and reference data. Adoption and utilization in routine clinical practice will also depend on the efficiency with which such measurements can be made.

With the goal of reserving the use of the term “spondylolisthesis” for <u>abnormal</u> sagittal plane positioning between vertebrae, the term “sagittal plane offset” (SPO) will be used generically for all measurements of the offset of one vertebra with respect to the adjacent vertebra. It is assumed that a range of SPO will be “normal” and SPO outside of that range can be labeled spondylolisthesis (either anterior or posterior). When SPO or disc height loss is pronounced, the reliability of current clinical assessment may be sufficient with current methods, but when it is more subtle, reliable reference data are needed as well as methods to account for confounding variables. It is also possible that new insights can be gained from using a continuous quantitative SPO metric in research studies. The challenge is to validate criteria for classifying “normal” versus “abnormal” SPO, that account for important variables and sources of error. Objective criteria for classifying disc heights or disc angles as normal vs abnormal are also poorly developed and additional reference data are needed, as evidenced by the lack of standardization in how SPO is measured and reported in research and clinical practice.

The rapid advancement of artificial intelligence/machine learning technology in the field of spine care will make near instant quantitative measurements of spondylolisthesis, disc heights, angles and other metrics possible^1–3^. That technology will require validated guidelines for how to optimally interpret these measurements. Once guidelines are validated, evidence can be generated to determine if and how automated quantitative measurements might improve outcomes and lower costs for patients with symptoms or treatment outcomes that may be related to abnormal SPO, disc heights or disc angles. The value of a quantitative diagnostic is in part determined by measurement errors, so understanding how to minimize sources of error in spondylolisthesis and disc measurements is a necessary prerequisite to effective clinical trials.

Multiple factors may influence SPO measurements. For example, SPO is dependent on patient positioning^4^. This may be due to the differences in spinal loading conditions between positions but may also be partially due to how SPO measurements are affected by disc angle and disc height. An objective metric that can account for variability in disc angle and disc height, while also accounting for differences between levels and sexes and other variables, may facilitate more precise diagnosis as well as allow for more sensitive detection of changes over time. If accurate measurement of SPO must account for multiple factors, it is possible this can be automated and accomplished algorithmically with reference data from a reliable analysis of a large population.

### Hypotheses

1. SPO is dependent on level (eg L1-L2 vs L4-L5), disc height and disc angle, as well as other variables.
2. If hypothesis 1 is true, then an objective metric to quantify SPO can be developed that accounts for: 1) the variability in disc angle due to patient positioning, 2) the normal variability in disc height between patients, 3) the variability in disc height between levels, and 4) the variability of normal SPO as it exists in asymptomatic volunteers at normal levels.
3. The above two hypotheses can be tested using the database of over 7,000 lumbar spine radiographs from the NHANES-II study, supplemented with a collection of flexion-extension radiographs to better understand the importance of disc angle.
4. SPO and disc metrics will depend in part on variability in radiographic projection and this phenomena can be understood by analysis of precisely calculated landmarks obtained from variable, digitally reconstructed radiographic projections.

### NHANES-II

Between 1976 and 1980, the 2nd National Health and Nutrition Examination Survey (NHANES-II) was conducted.^5^ This was a nationwide probability sample to document the health status of the United States. Justified by the prevalence and societal impact of back pain, approximately 7,000 lateral lumbar spine X-rays were collected as part of this survey. One lateral lumbar X-ray was obtained for each participant. Demographic, anthropometric, health, and medical history data were also collected. NHANES-II may thereby be useful to establish normative reference data for use in clinical practice and research studies.

## Materials and Methods

The NHANES-II images and data were obtained through public access^2^. A previously validated series of neural networks and coded logic (Spine CAMP^TM^, Medical Metrics, Inc., Houston, TX) were used to automatically obtain four landmarks for each vertebral body from L1 to S1 (Supplemental Appendix 1, Figure 1). Although the NHANES-II lumbar X-rays almost always included up to T10 in the field-of-view, the neural networks were only trained to recognize L1 to S1. The series of neural networks include quality control networks to assure the X-ray was a lateral lumbar view, manipulate the image if needed so upper vertebrae are toward the top of the X-ray, assure spinous processes toward the left side of the image, and assure that bone appears whiter than air. Neural networks and coded logic were then used to segment the bone, find individual vertebrae, label the vertebrae, find the four corners of each vertebra, and finally refrain from reporting landmark coordinates when the confidence of the networks was too low. In addition, an anomaly detection algorithm was used to identify likely errors in landmark placement. Those levels were excluded from the data analysis. All the neural networks were trained with over twenty thousand lateral X-rays where analysts had previously digitized standardized landmarks. The NHANES-II images were not used in training the neural networks. The analysts had used Quantitative Motion Analysis software (QMA®, Medical Metrics, Inc, Houston, TX) to place the landmarks. The QMA® software has previously been validated. ^6–11^ QMA® has been used to produce data reported in over 170 peer-reviewed spine studies (references available upon request). Vertebral labeling in the data used to train the neural networks was based on identifying the vertebra most characteristic of S1 and then labeling other vertebrae relative to S1.

**Figure 1:**
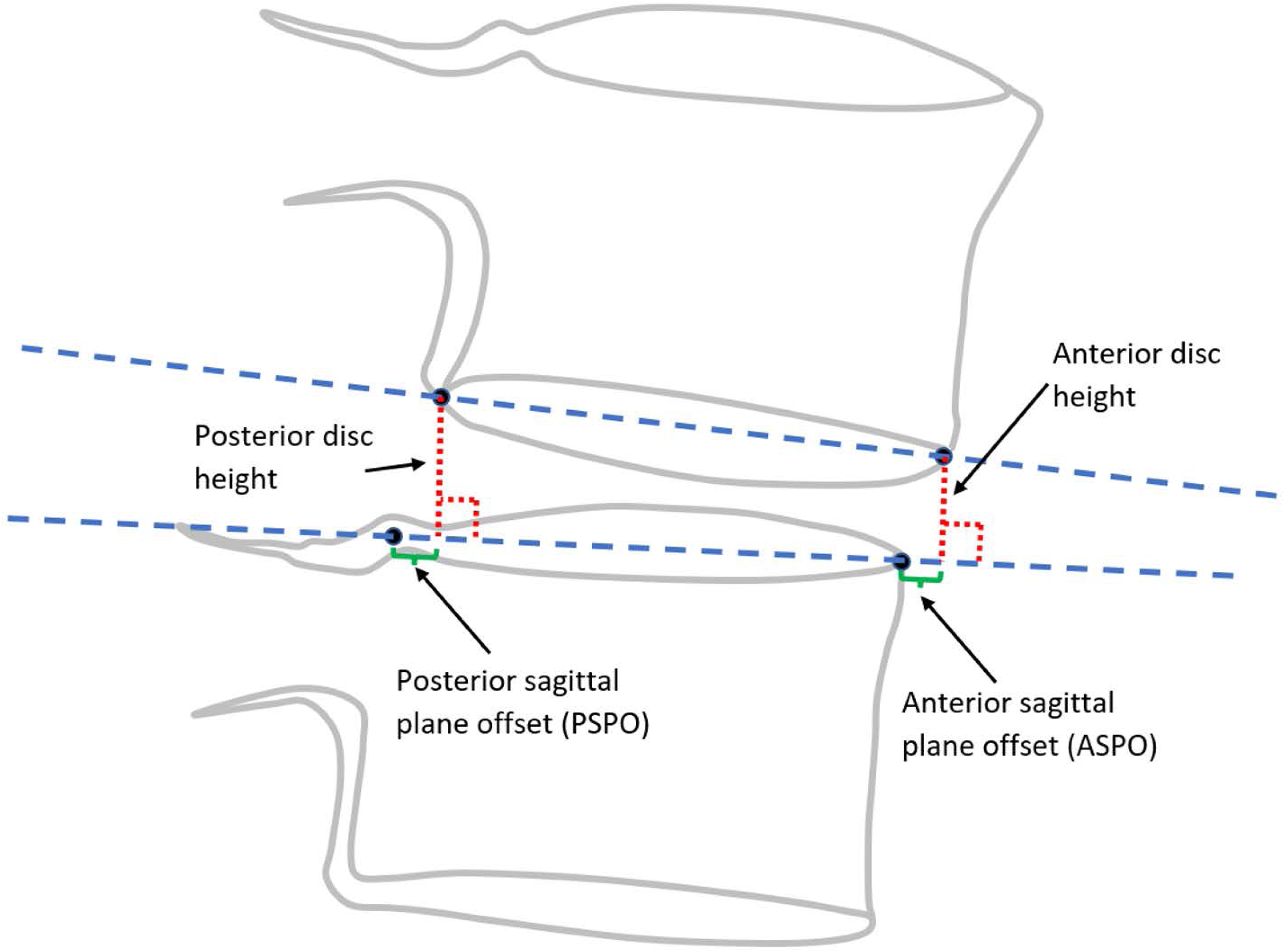
Disc height and SPO measurements using method 1.

Details of landmark placement are provided in Supplemental Appendix 1, since all SPO and disc measurements will be dependent on exactly how the landmarks are placed^12^. Vertebral morphology calculated from the landmarks has been previously reported.^13^

The following nine metrics were calculated from the landmarks for each intervertebral level from L1-L2 to L5-S1, using one of two methods:

- Method 1

- Anterior disc height (ADH)
- Posterior disc height (PDH)
- Disc angle (DA)
- Anterior SPO (ASPO)
- Posterior SPO (PSPO)
- Method 2

- Ventral disc height (VDH)
- Dorsal disc height (DDH)
- Mid-plane angle (MPA)
- Centroid SPO (CSPO)

The average disc height was also calculated from ADH and PDH as that may be of value in some applications. The disc area was also calculated (in units of endplate width squared) from the four landmarks using the shoelace algorithm, as this may be of interest in some applications.

Method 1 metrics are illustrated in Figure. Method 2 metrics were all first described by Frobin et al and are illustrated in Figures 2 and 3.^14,15^ Method 2 was developed by Frobin et al with the goal of minimizing the influence of radiographic distortion on measurements. The ventral and dorsal terminology used by Frobin et al is retained in this paper to help distinguish between height metrics calculated using method 1 versus method 2. Since substantial variability can occur in radiographic magnification^16–18^, and since there was no scaling device in the NHANES-II X-rays, all SPO and disc height metrics were calculated in units of % endplate width, using the superior endplate of the inferior vertebra. Note that interpretation of the sign of spondylolisthesis is based on local anatomy and not global anatomy. A positive ASPO informs that the anterior inferior corner of the superior vertebra is anterior to the anterior superior corner of the inferior vertebra. A positive PSPO informs that the posterior inferior corner of the superior vertebra is posterior to the posterior superior corner of the inferior vertebra. A negative DA or MPA informs that the ADH (or VDH) is smaller than the PDH (or DDH).

**Figure 2:**
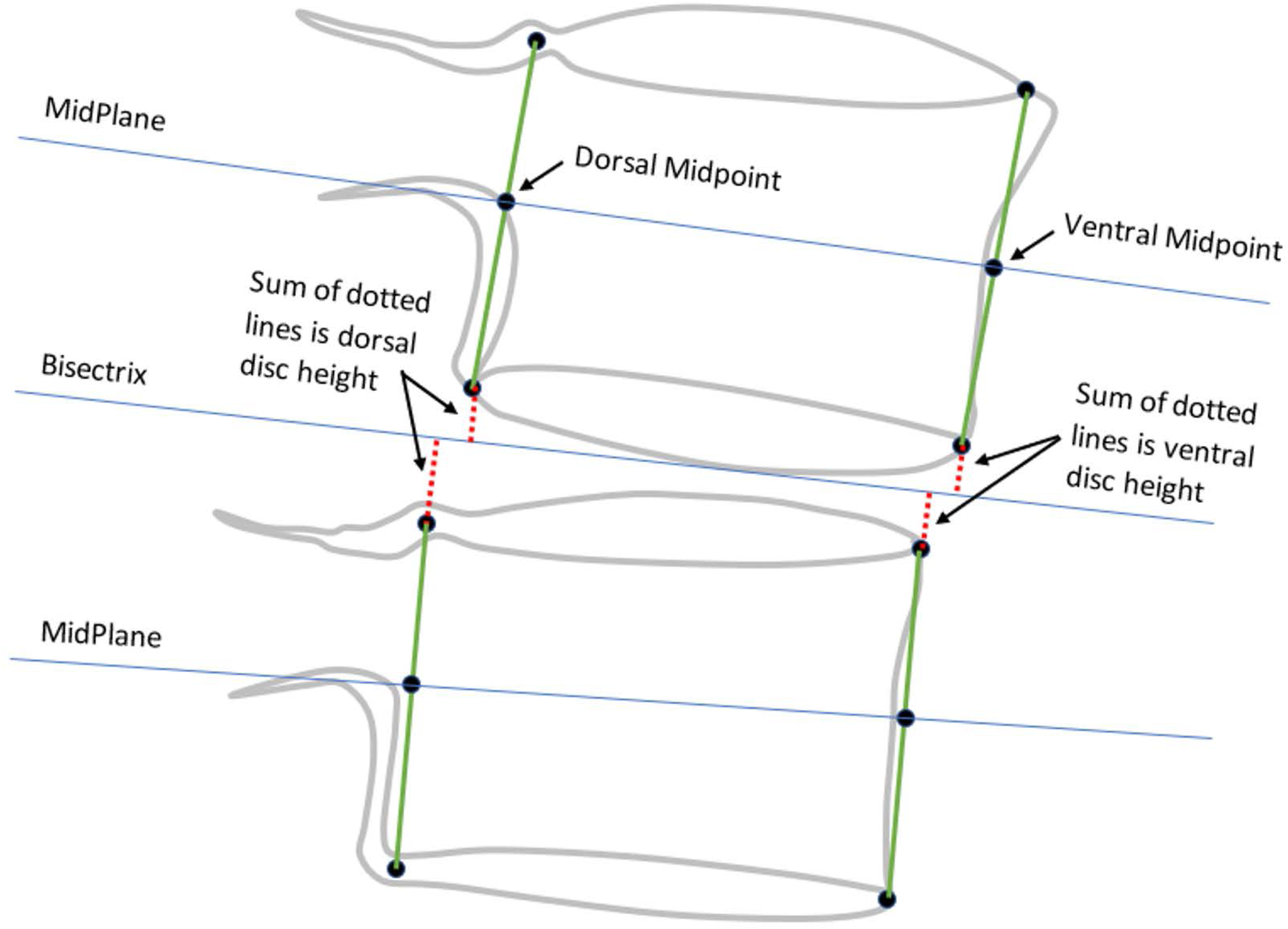
Disc height and SPO measurements using method 2 (adapted from Frobin et al^14^).

**Figure 3:**
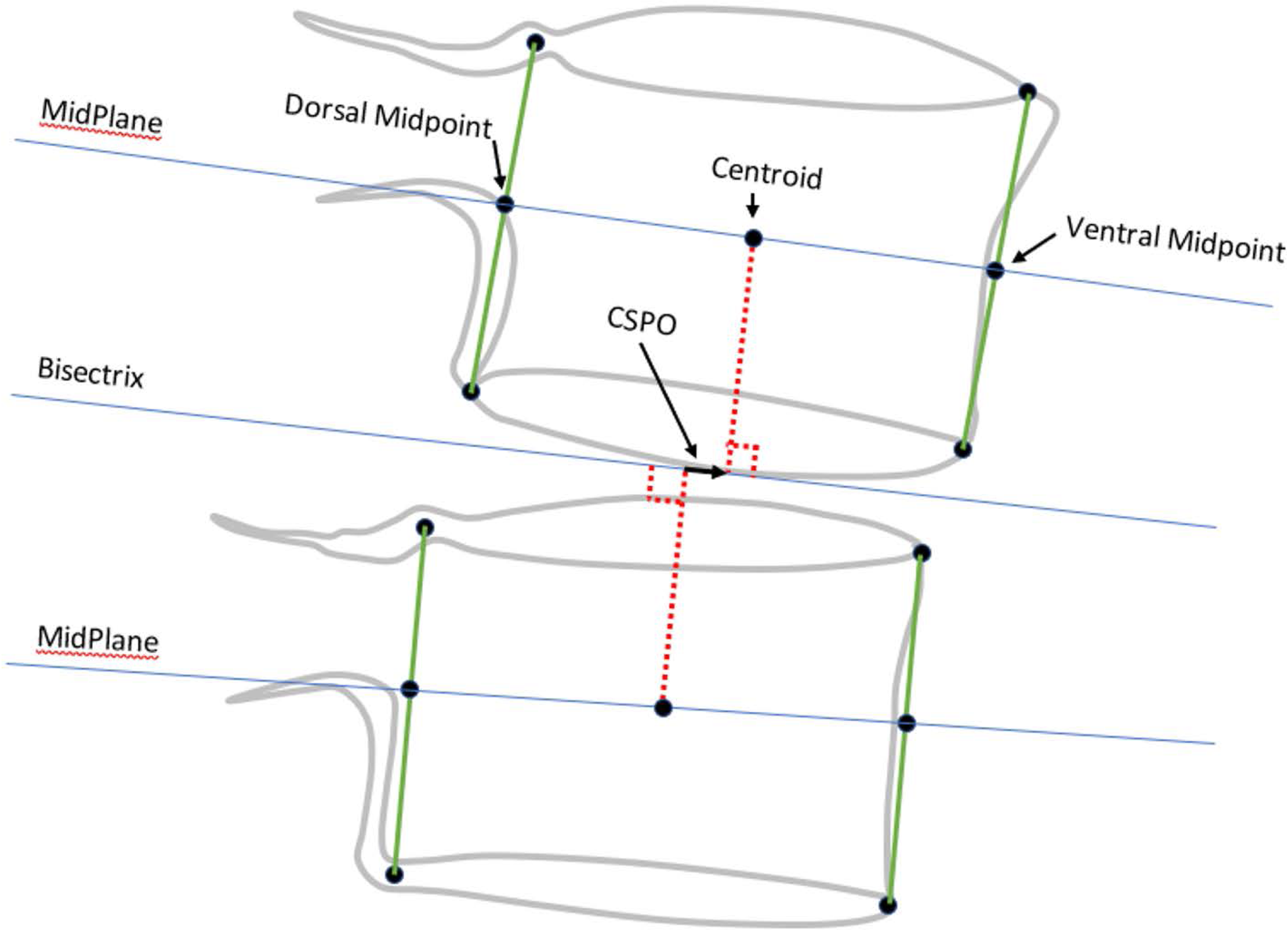
Method used to measure Centroid Sagittal Plane Offset (CSPO), adapted from Frobin et al.^15^. The dotted lines connect the centroids to the disc bisectrix and are perpendicular to the bisectrix. CSPO therefore measures the offset between the vertebral centroids along the disc bisectrix.

With the goal of establishing normal SPO, disc heights, and disc angles, abnormalities in the NHANES-II database were trimmed based on the distribution of the data. The goal was to achieve normally distributed data that describe healthy disc metrics and SPO, so outliers were trimmed based on percentiles in the distributions. Multivariate analysis of variance was used to understand associations between the SPO and disc metrics, and age, sex, BMI and other variables. All statistical analysis was completed using Stata ver 15 (College Station, TX).

Since all the NHANES-II radiographs were obtained with the subject side-lying, there was generally not a lot of variation in the flexion angle relative to what can occur in clinical practice. To better understand the effect of disc angle and disc heights on SPO, flexion-extension radiographs from a prior study^19^ were also analyzed to obtain a larger range of disc angles. Regression analysis with post-estimation calculations was used to establish coefficients for equations that allow for predicting normal SPO from the flexion-extension radiographs based on the most predictive variables, excluding the outliers. This is described in Supplemental Appendix 2.

It is known that variability in radiographic projection can affect measurements of the spine.^15,20^ To help appreciate the magnitude of error that can occur in the nine SPO and disc metrics, simulated X-rays were used that were generated with precise knowledge of the true location of the landmarks on the X-ray. This experiment is described in Supplemental Appendix 3.

## Results

### Summary of Data Analyzed

SPO and disc metrics were obtained for 35,446 levels from 7,358 NHANES-II subjects who were 25 to 74 years old. It took approximately 9 hrs on a dual CPU, 4 GPU (Nvidia T4) production server to generate the landmarks for all X-rays, that were then used to calculate the disc and SPO metrics. Data were not produced for 1300 levels (3.5% of total possible levels) due to issues detected by the neural networks and coded logic. An additional 44 levels (0.12%) were excluded by anomaly detection. The age and body size data are summarized in Table 1. Older ages are disproportionately represented (Figure 4).

**Figure 4:**
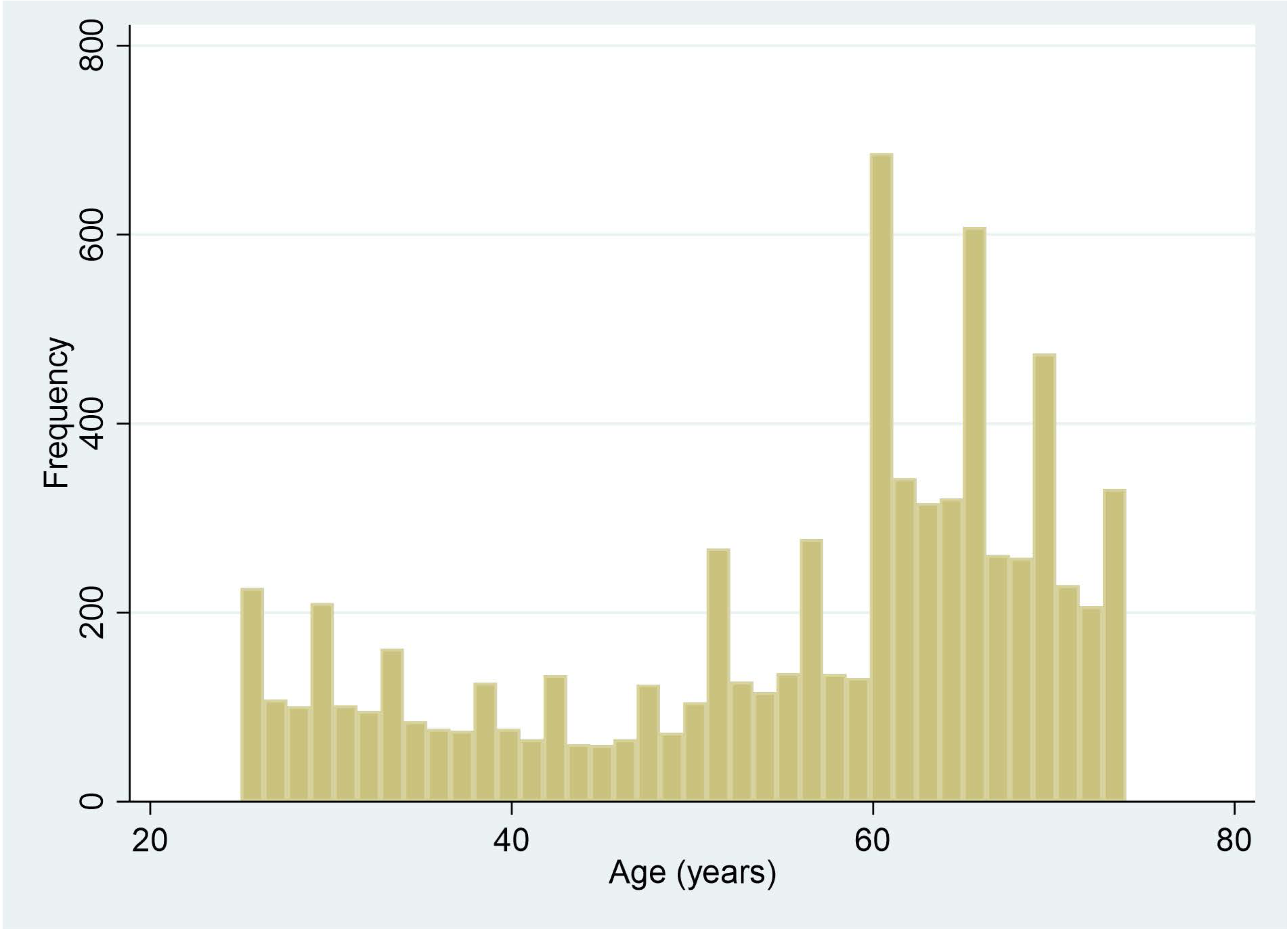
Distribution of ages in the NHANES-II study.

**Table 1:** Sample size, average age and [SD], and average BMI (kg/m2) and [SD] for the NHANES-II subjects that were analyzed.

| Sex | N | Age | BMI |
| --- | --- | --- | --- |
| Male | 4,552 | 50.9[15.3] | 25.5[4.0] |
| Female | 2,806 | 63.3[6.4] | 26.4[5.5] |

### Correlations between disc and SPO metrics

The method 1 and method 2 disc and SPO metrics were significantly correlated with each other (Pearson correlation coefficients between each pair of variables, for the L3L4 level, are provided in Table 2). Correlations were nearly perfect between anterior disc height and ventral disc height, between posterior disc height and dorsal disc height, and between average disc height and disc area. Disc angle (method 1) and mid-plane disc angle (method 2) were also strongly correlated. Note that ASPO and CSPO are positively correlated while PSPO and CSPO are negatively correlated since PSPO quantifies the amount of translation of the posterior-inferior corner of the superior vertebra into the spinal canal.

**Table 2:** Pearson correlation coefficients between each pair of variables, for the L3L4 level, including data for all subjects. Every correlation coefficient is statistically significant in this table (P<0.0001). Similar results were found at all other levels.

|  | ADH | PDH | Avg<br>DH | DA | ASPO | PSPO | VDH | DDH | MPA | CSPO | Disc<br>AREA |
| --- | --- | --- | --- | --- | --- | --- | --- | --- | --- | --- | --- |
| ADH | 1 |  |  |  |  |  |  |  |  |  |  |
| PDH | 0.46 | 1 |  |  |  |  |  |  |  |  |  |
| AvgDH | 0.90 | 0.80 | 1 |  |  |  |  |  |  |  |  |
| DA | 0.70 | -0.30 | 0.32 | 1 |  |  |  |  |  |  |  |
| ASPO | -0.31 | 0.20 | -0.11 | -0.52 | 1 |  |  |  |  |  |  |
| PSPO | 0.29 | -0.07 | 0.16 | 0.34 | -0.74 | 1 |  |  |  |  |  |
| VDH | 0.99 | 0.43 | 0.87 | 0.73 | -0.39 | 0.27 | 1 |  |  |  |  |
| DDH | 0.48 | 0.99 | 0.81 | -0.27 | 0.13 | -0.05 | 0.46 | 1 |  |  |  |
| MPA | 0.53 | -0.32 | 0.20 | 0.82 | -0.36 | 0.19 | 0.55 | -0.30 | 1 |  |  |
| CSPO | -0.15 | 0.11 | -0.05 | -0.26 | 0.80 | -0.83 | -0.18 | 0.07 | -0.17 | 1 |  |
| Disc<br>AREA | 0.91 | 0.78 | 1.00 | 0.35 | -0.13 | 0.23 | 0.88 | 0.79 | 0.21 | -0.09 | 1 |

### Establishing normal disc and SPO metrics

With the goal of using the NHANES-II X-rays to develop reference data that can define “normal” disc heights, angles, and SPO, outliers were trimmed based on percentiles. Initial assessment of the distributions of disc metrics, including all NHANES-II subjects, revealed that every metric was significantly skewed with significant kurtosis (p<0.0001 for both skewness and kurtosis, for every metric at almost every level). One set of trimming criteria (such as excluding the bottom and top 25%) was not used for all metrics, because the distribution of data post-trimming was clearly not normal for all metrics. This is understandable, since for example, disc height loss from degeneration is far more likely than abnormal disc space widening. If too much of the tails of the entire dataset were removed for any metric, the tails of the distribution were clearly truncated (not a gaussian distribution) and/or the distribution was skewed. Table 3 describes the criteria used to trim outliers for each of the variables. The proportions of the upper and lower tails that were trimmed per Table 3 were selected to achieve skewness close to 0 and kurtosis close to 3 (ideal for a normal distribution), although a formal optimization scheme was not used.

**Table 3:** criteria for identifying and trimming outliers in the data.

| Metric | Acronym | Lower Proportion<br>Removed | Upper Proportion<br>Removed |
| --- | --- | --- | --- |
| Method 1 |  |  |  |
| Anterior Disc Height | ADH | 10 | 5 |
| Posterior Disc Height | PDH | 10 | 5 |
| Disc Angle | DA | 10 | 10 |
| Anterior Sagittal Plane Offset | ASPO | 5 | 5 |
| Posterior Sagittal Plane Offset | PSPO | 5 | 5 |
| Method 2 |  |  |  |
| Ventral Disc Height | VDH | 10 | 5 |
| Dorsal Disc Height | DDH | 10 | 5 |
| MidPlane Disc Angle | MPA | 10 | 10 |
| Centroid Sagittal Plane Offset | CSPO | 5 | 5 |

Supplemental Appendix 1, Tables 1 to 10, provide descriptive statistics for the NHANES-II lumbar SPO and disc metrics, before and after trimming the data. Note that trimming: 1) eliminates approximately half of the data, since if any of the nine metrics was an outlier (based on Table 3), all data for that level was excluded; 2) does not substantially change the mean or median; 3) substantially lowers the coefficient of variation; 4) brings skewness close to 0 and kurtosis closer to 3.

### Analysis of sources of variance in SPO metrics

Based on multivariate analysis of variance (ANOVA), after excluding trimmed data (per criteria in Table 3), ASPO and PSPO (method 1) were significantly dependent on level, anterior and posterior disc heights, disc angle, age, and BMI (P<0.0001 for all variables). Sex, race, and nation of origin were not significant for ASPO (P>0.78 for all three). Based on the F-value, disc angle had the strongest association with ASPO, followed by anterior and posterior disc height. The F-value for level and age was an order of magnitude less than for disc angle and disc heights. Sex had a significant association with PSPO, though the F-value was very small. Based on F-values, disc heights and disc angle had the strongest association with PSPO with the F-value for age and level an order of magnitude lower. However, the ANOVA R^2^ was 0.56 for ASPO and 0.44 for PSPO, and that would likely not be sufficient to facilitate reliably predicting normal SPO from disc angle and disc height. Too much of the variability would be left unexplained.

Based on multivariate analysis of variance (ANOVA), after excluding abnormal measurements (per criteria in Table 3), CSPO (method 2) was significantly dependent on level, ventral and dorsal disc heights, age, BMI (P<0.0001), sex (P=0.0002), and on midplane angle(P=0.006). Race and nation of origin were not significant (P>0.83). Intervertebral level was over an order of magnitude more important than any other variable. The F-value for midplane angle, BMI and sex were small. Including only level, R^2^ = 0.76 and this was only slightly lower than when all variables were included (R^2^ = 0.77). Thus, with CSPO, only level is needed to predict normal CSPO, and almost 80% of the variability in CSPO can be explained by level.

### Applicability of NHANES-II SPO reference data to flexion-extension X-rays

Supplemental Appendix 2 describes the results of applying the NHANES-II SPO reference data to flexion-extension studies of asymptomatic volunteers. Applying the normative data from the NHANES-II study (where all subjects were imaged in a side-lying position) to 373 flexion-extension radiographs (where subjects were upright), 45% of the flexion and extension radiographs have standardized ASPO metrics >2 or < −2 Std Dev from average normal ASPO. Since 45% of asymptomatic levels can’t have abnormal SPO, it is clear that application NHANES-II normative SPO data to flexion and extension X-rays is NOT appropriate.

Supplemental appendix 2 also describes development of an equation that can quantify SPO, accounting for disc heights and disc angles, in the presence of flexion and extension. That equation provides the spondylolisthesis index (SI) which is an estimate of the deviation from normal ASPO that accounts for disc heights and disc angle, and can be applied to neutral X-rays as well as flexion-extension X-rays. Applying that equation to the NHANES-II data, 16% of levels had a SI > 2 and 7% had a SI > 3. Most of the abnormalities were at L4-L5 and L5-S1. The SI and the normalized ASPO metric were linearly correlated (R^2^=0.64)

### Prevalence of abnormalities in the NHANES-II lumbar spine radiographs

Using the post-trimming data in tables 1 to 10 of Supplemental Appendix 1, each measurement in the NHANES-II study was classified as “normal” or “abnormal” (eg ventral disc heights in the lowest 10^th^ percentile were classified as abnormal). The prevalence of abnormalities in the NHANES-II data is provided in Supplementary Appendix 1, Tables 11 to 16. Prevalence was calculated using the standardized (“z”) version of the metrics to simplify interpretation of the data. Both the prevalence of Z scores > 2 and > 3 are provided.

### Associations with Back Pain

The subjects in the NHANES II study were asked the question: “Have you ever had pain in your back on most days for at least two weeks”. It is challenging to test for associations between the response to the question and the disc and SPO metrics since it is not clear what level(s) in the spine might be associated with symptoms. Nevertheless, the maximum absolute standardized ASPO, PSPO, and CSPO were calculated for each spine. Logistic regression documents a significant association between maximum absolute standardized deviation from normal SPO and whether the subject reported ever having back pain, P<0.05. However, the odds ratios were close to 1 (1.04 for ASPO and PSPO, and 1.05 for CSPO). Thus, although an association between SPO and back pain can be detected using the NHANES-II data, it is weak. Similarly, the minimum standardized anterior and posterior disc heights were calculated for each subject. Minimum ventral and dorsal disc heights were not calculated since these are nearly perfectly correlated with anterior and posterior disc heights. Both minimum standardized anterior and minimum standardized posterior disc heights were significantly associated with whether the subject reported ever having back pain (P<0.0001). The odds ratios (0.87 for standardized ADH, and 0.85 for standardized PDH) suggest a stronger association between back pain and loss in disc height than found for SPO. The odds ratios from logistic regressions between maximum absolute standardized disc angle and maximum absolute standardized mid-plane angle were also significant, documenting that deviation at any level within a spine from normal disc angle (P=0.001, OR=1.09) or from normal midplane angle (P=0.015, OR 1.07) can be associated with back pain.

## Discussion

A standardized definition of normal SPO and disc metrics, that can be used across research studies and in clinical practice may facilitate advancements in spine care. Analysis of the NHANES data were used to define normal SPO and other disc metrics in an attempt to achieve the goal of a standardized definition. The landmarks used to calculate the metrics were produced using a proprietary network of neural networks and coded logic. However, the landmarks are intended to be equivalent to landmarks produced for many prior research studies. Artificial intelligence has been used to produce vertebral landmarks in many prior studies.^(62–67)^ It is expected that the landmark placement obtained for the NHANES-II radiographs can be reproduced by other methods and the NHANES-II reference data in Supplemental Appendix 1 will thereby by useful in other studies where landmarks are obtained with alternative methods.

Further research is needed to determine how to effectively use SPO and other metrics in research and clinical practice. The NHANES-II SPO and disc metrics also document that while some strong correlations exist between a few of these metrics, other metrics are not as strongly correlated supporting that most of the metrics are, in part, detecting unique aspects of disc height, angle, and SPO. The NHANES-II data also support hypothesis 1 in showing that quantitative measurement of SPO are significantly correlated with disc heights and disc angles, though a lot of variability in SPO is not explained by disc heights and disc angles. A prediction equation developed and described in Supplemental Appendix 2 provides an improved method to account for disc heights and disc angles when predicting what should be “normal” SPO for a level. That equation explains more of the variability in SPO.

Multiple SPO and disc metrics were investigated. In Figure 1, note that the method 1 anterior and posterior spondylolisthesis measurements are local measurements that quantify the position of a specific corner of the superior vertebra relative to a specific corner of the inferior vertebra, whereas the method 2 displacement of the centroid (Figure 3) is a more generic measurement of the position of the superior vertebra relative to the disc bisectrix between superior and inferior vertebrae. One goal of method 2 was to reduce the effects of radiographic distortion, as explained by Frobin et al.^14^ The NHANES-II data support that CSPO may be the best metric to use for assessing overall SPO. ASPO and PSPO may prove more effective if the local effects of SPO are important. Carefully contemplating Figures 1 to 3, with respect to how loss of disc height or a change in disc angle might change the SPO measurements, helps to appreciate the hypothesis that disc height and disc angle could change SPO measurements. The research and clinical efficacy of the method 1 versus method 2 SPO measurements remains to be determined.

A primary goal was to use the NHANES-II X-rays to establish normal sagittal plane SPO such that abnormal SPO (spondylolisthesis) can be identified and quantified. There currently is no well-validated “gold standard” metric for abnormal versus normal SPO. Insufficient resources prevented manual clinician assessment of all NHANES-II X-rays. Even if manual assessments of disc height, angles and SPO had been obtained, error due to observer variability would be expected. ^11,21–23^ Thus, analysis without a gold standard was required. This prevented traditional sensitivity/specificity analysis.

Expert consensus is that a standing neutral-lateral radiograph should be used for diagnosis of sagittal plane spondylolisthesis, although it can also be measured in flexion and extension.^24,25,26^ There is limited published evidence about how flexion and extension may affect the assessment of spondylolisthesis. Considering the Pearson correlation coefficients in Table 2, all three SPO metrics were significantly correlated with disc angle. A variable amount of disc height loss can also coexist with spondylolisthesis and there is limited evidence how variability in disc heights effects the assessment of spondylolisthesis. The data from the NHANES-II study, as well as the analysis reported in Supplemental Appendix 2 may help to establish comprehensive and standardized SPO assessment.

One of the most compelling pieces of evidence in support of the need for considering disc heights and angles when assessing SPO was observed when applying the NHANES-II SPO reference data to flexion-extension X-rays (Supplemental Appendix 2). Very large differences in the NHANES-II based standardized ASPO, PSPO, and CSPO metrics were observed between the flexion and extension X-rays for each subject. The NHANES-II standardized ASPO, PSPO, and CSPO metrics account for the relatively low variability that occurs in a collection of X-rays that were all obtained in the lateral decubitus position. The NHNAES-II SPO data are clearly not applicable to X-rays taken in a flexed or extended position, unless the thresholds used to classify a level as normal vs abnormal are set to a high level. In Supplemental Appendix 2 a “Spondylolisthesis Index” (SI) is developed that accounts for variability in disc heights and disc angles. SI is also reported in units of deviation from average normal at radiographically normal levels in asymptomatic volunteers.

The experiment using simulated X-rays, described in Supplemental Appendix 3, documents that some error in the disc height, angle and SPO measurements occurs due to variability in radiographic projections. As an example, the median error in measuring ADH from a radiograph was 0.01 std dev from average normal ADH. This is the typical variability expected only due to radiographic projection. However, for highly out-of-plane imaging of a disc space, errors can approach 1 std dev solely due to the poorly imaged level. Solutions to this issue with lateral spine radiographs include protocols to optimize imaging of each level and possibly neural networks trained to correct for out-of-plane imaging.

There are multiple approaches to both imaging and measuring spondylolisthesis and this must be considered when attempting to compare data between studies.^4,27^ There is as yet no validated consensus for how best to measure and interpret spondylolisthesis. The criteria for interpreting a spondylolisthesis measurement must be specific to how the measurement is made to avoid incorrect classification of spondylolisthesis as normal versus abnormal.^27^ Reliability of the measurements depends on the method used.^23,27^ Measurement error has been documented. ^28–33^. In one study, spondylolisthesis was measured with interclass correlation coefficients that were generally >= 0.8, with better results when measured on a workstation allowing zooming in on the anatomy.^34^ Interclass correlation coefficients and other metrics can document reproducibility, but accuracy is also important. Finally, optimizing diagnosis of spondylolisthesis is dependent on understanding the variables that affect interpretation of the measurements.

Clinical studies of treatments for symptomatic spondylolisthesis will typically report that patients had spondylolisthesis but do not give any details about how the assessment was made, other than perhaps a threshold amount of SPO (eg Austevoll et al^35^). A threshold level of 3 mm is commonly used. However, it is necessary to correct for image magnification when measuring SPO in millimeters, since magnification can be highly variable. ^16^ PSPO can normally vary by over 8.56 % endplate width based on trimmed data in Supplemental Appendix 1 for the L4L5 level. Using 35 mm as a typical endplate width, then approximately 3 mm of PSPO can be within normal limits. With high magnification, 3 mm of SPO would likely not be abnormal. To avoid errors when radiographic magnification is unknown, it may be preferable to measure SPO as % endplate width and use appropriate reference data.

The SPO measurement method that will prove of greatest clinical benefit has yet to be determined. Sagittal plane displacement of the centroid of the vertebral body (Figure 3) may be important when the displacement of the entire superior vertebral body relative to the inferior vertebra is prognostic. A more localized measurement, such as the posterior-inferior corner of the superior vertebra relative to posterior-superior corner of the inferior vertebra (PSPO) might be of greater significance when inflammation of the tissue within the neural foramina immediately adjacent to the vertebral corner is a source of symptoms. The descriptive statistics in Supplemental Appendix 1 document that, excluding outliers, ASPO, PSPO, or CSPO is level dependent and rarely exceeds 14% endplate width.

In addition to measurement of the magnitude of SPO, there are different aspects of spondylolisthesis that may be clinically important ^36,37^. The degenerative form of spondylolisthesis is associated with arthritis and hypermobility of the facet joints.^38–40^ It has also been shown that hypoplasia of L5 can occur and this can create a false impression of spondylolisthesis.^41–43^ Vertebral morphology data from NHANES-II can help to identify hypoplasia and other abnormalities.{Hipp, 2022 #10614} It may also be important to interpret SPO with respect to factors such as pelvic incidence.^31,44^ An alternative classification system was described by Marchetti-Bartolozzi.^45^ The relative importance of SPO magnitude versus the other aspects of classifying spondylolisthesis remains to be determined. It is likely that with sufficient training material, artificial intelligence can be trained to objectively apply different spondylolisthesis classification systems that are currently based primarily on subjective assessment. That could supplement basic quantitative metrics.

Multiple prior studies help with assessing the prevalence of abnormal SPO in the NHANES-II as documented in Supplemental Appendix 1. Kalichman et al found a prevalence of 21% in the Framingham heart study, with the prevalence increasing with age.^46^ That is higher than in the NHANES-II study, but the population sampling is different. There is some evidence of an association with occupational exposure.^47^ There are relevant data collected in the NHANES-II study that could be use to test this and similar associations. Importantly with respect to the spondylolisthesis index, Cehn et al found that the disc height is a predictor of spondylolisthesis, as well as the ratio of posterior to anterior vertebral body height.^48^ It would be interesting to know what might be learned if all these studies were repeated with objective and standardized SPO metrics.

Although SPO was a primary focus of the study, a lot of disc height and disc angle data were generated that may prove useful. The disc height data in particular may help with assessing for loss in disc height, which may prove important in multiple scenarios, such as identifying candidates for biologic disc treatments where the treatment may be more effective in treating the early stages of disc degeneration. The association of disc height with age in the NHANES-II study was significant, though weak. Age explained < 1% of the variation in ADH and VDH, and 4% of the variation in PDH and DDH (P<0.0001). Whereas several authors have reported an association of disc height with age (either disc height increases with age or decreases with age, depending on the study), the strength of the association is very important to consider. ^49–54^ It is also possible that reporting disc height in units of percent endplate width minimize the differences between sexes compared to studies showing a difference in disc height between males and females when disc height is measured in millimeters. ^53,54^

Despite that publications and spine fusion coverage policies infer interchangeability of spondylolisthesis with instability^55^, this has been proven in several studies NOT to be true.^56–59^ There is no way to address this issue with the NHANES-II data, in large part due to the lack of validated diagnostic metrics for instability.^60–62^ Critical assessment of the available evidence base supports that the clinical importance of, and optimal treatment for spondylolisthesis is not fully understood.^63–66^ Assessing whether the spondylolisthesis is static or dynamic is likely very important^67,68^. That could not be assessed using the NHANES-II data. Development of the evidence base will likely be facilitated by objective and standardized measurements that are consistently used across research studies.

Limitations of this study include:

1. Poor representation of some races, nations of origin, and sex in some age groups. In particular, females are under-represented. By design of the NHANES-II study, no lumbar X-rays were taken for pregnant women or women under 50.^69^ An additional limitation relates to the data on race and nation of origin recorded in the NHANES-II data. 86.9% of NHANES-II subjects were “white”, 11.2% “black” and the rest “other”. A more uniform representation of races would likely be needed to fully understand the importance of race. The same is true for the “nation of origin” data in NHANES-II. *NOTE: the language in quotes is as used in the NHANES-II documentation:* 74.1% of subjects are classified as “OTHER EUROPE SUCH AS GERMAN, FRENCH, ENGLISH, IRISH”, 10.8% classified as “BLACK, NEGRO OR AFRO-AMERICAN”, and the rest distributed in 1 of 11 other classifications. It would be valuable to repeat this analysis on X-rays that better represent races and nations of origin that are not well represented in the NHANES-II data. The ages of the subjects were also biased toward older ages (Figure 4).
2. NHANES-II lumbar spine X-rays were obtained side-lying. Disc metrics in Appendix 1 may not be directly and precisely applicable to X-rays taken in other positions. Using all of the NHANES data, the L1-S1 angle was 51.4 (std Dev 12.7). Table 4 provides data from three prior studies reporting L1-S1 angles measured from X-rays obtained with subjects standing.^70–72^ The lordosis data from the NHANES-II study is very similar to lordosis measured from standing X-rays in three other studies. Figure 5 shows mid-plane angles from the NHANES-II study, excluding levels where any disc or SPO metric was abnormal, comparted to mid-plane angles reported by Frobin et al from X-rays obtained with subjects standing.^14^ There are some differences, but also some similarities. These comparative data can help in deciding whether to use NHANES-II reference data with standing or other X-rays. With respect to external validity of the disc height measurements, comparable reference data were hard to find. There are multiple publications reporting lumbar intervertebral disc height reference data in units of millimeters, frequently from MRI or CT exams. Since a scaling device was not included in the NHANES-II X-rays, these prior publications cannot be used as comparative data. Data for a large collection of radiographs obtained in other positions and analyzed using the same methods would be required to understand applicability of the NHANES-II data to other protocols for obtaining lateral lumbar spine X-rays. It is likely that the average disc height data will be the most universally applicable of the NHANES-II reference data, since average disc height should minimize minor differences due to positioning.
3. The accuracy study using simulated X-rays (Supplemental Appendix 3) revealed potentially large errors in disc and SPO metrics when the radiographic projection is poor. It would be valuable to establish a neural network or other method to either make a correction in the metrics when large out-of-plane imaging is encountered, or to abstain from reporting data when vertebrae are poorly imaged. Nevertheless, a certain level of error must be expected in SPO and other metrics. This may be a particularly significant issue with large amounts of frontal plane spinal curvature. This study does not provide clear guidance on this issue and additional data are needed. Caution should be used when interpreting disc metrics and SPO from lateral X-rays where the vertebrae are poorly imaged.
4. One of the greatest challenges was in trimming the data such that only truly normal discs were used to define “normal” disc and SPO metrics. Abnormal disc heights, angles, and SPO can be expected in the NHANES-II study (since there was no attempt to exclude spine abnormalities). It was assumed that normal disc heights, disc angles, and SPO would have a gaussian distribution. It was also assumed that discs with abnormal heights, angles, or SPO, due to degeneration, trauma, genetics or other reasons would be in the tails of the distributions when all NHANES-II X-rays are combined. Even in healthy discs, a range of disc heights, angles, and SPO would be expected due to genetic differences between individuals (and possibly nutritional history and other variables). In addition, in a normal disc, the resting position of the vertebrae, when the radiograph was obtained, would be expected to be in the neutral zone. Within the neutral zone, little force is required to produce sagittal plane movements.^73,74^. It can be hypothesized that the amount of SPO may change slightly, but remain within the neutral zone, every time a person assumes a “neutral” position. That would be expected to contribute to a normal distribution of SPO within a population. There is currently no gold standard method that is validated for classifying discs as normal versus abnormal. MRI exams were not available. In the current study, an attempt was made to trim data so as to achieve a gaussian distribution. This approach has been used by other authors.^75–77^ A better strategy may be possible for assuring that only truly normal discs are used to define normative reference data. For example, a formal optimization scheme might be used to obtain a gaussian of a distribution, though justification for such an optimization is not well developed.
5. Although Supplemental Appendix 1 provides data to calculate disc and SPO metrics as number of Std Dev from average normal, a clinically meaningful threshold must be validated for use with these reference data that can classify a metric as normal vs abnormal. Even though a metric that is two Std Dev from average would be technically outside of the 95% confidence interval used to define “normal”, that may not be clinically significant. Until well-designed clinical trials are completed, the threshold level for the standardized score that is clinically efficacious will not be known.

**Figure 5:**
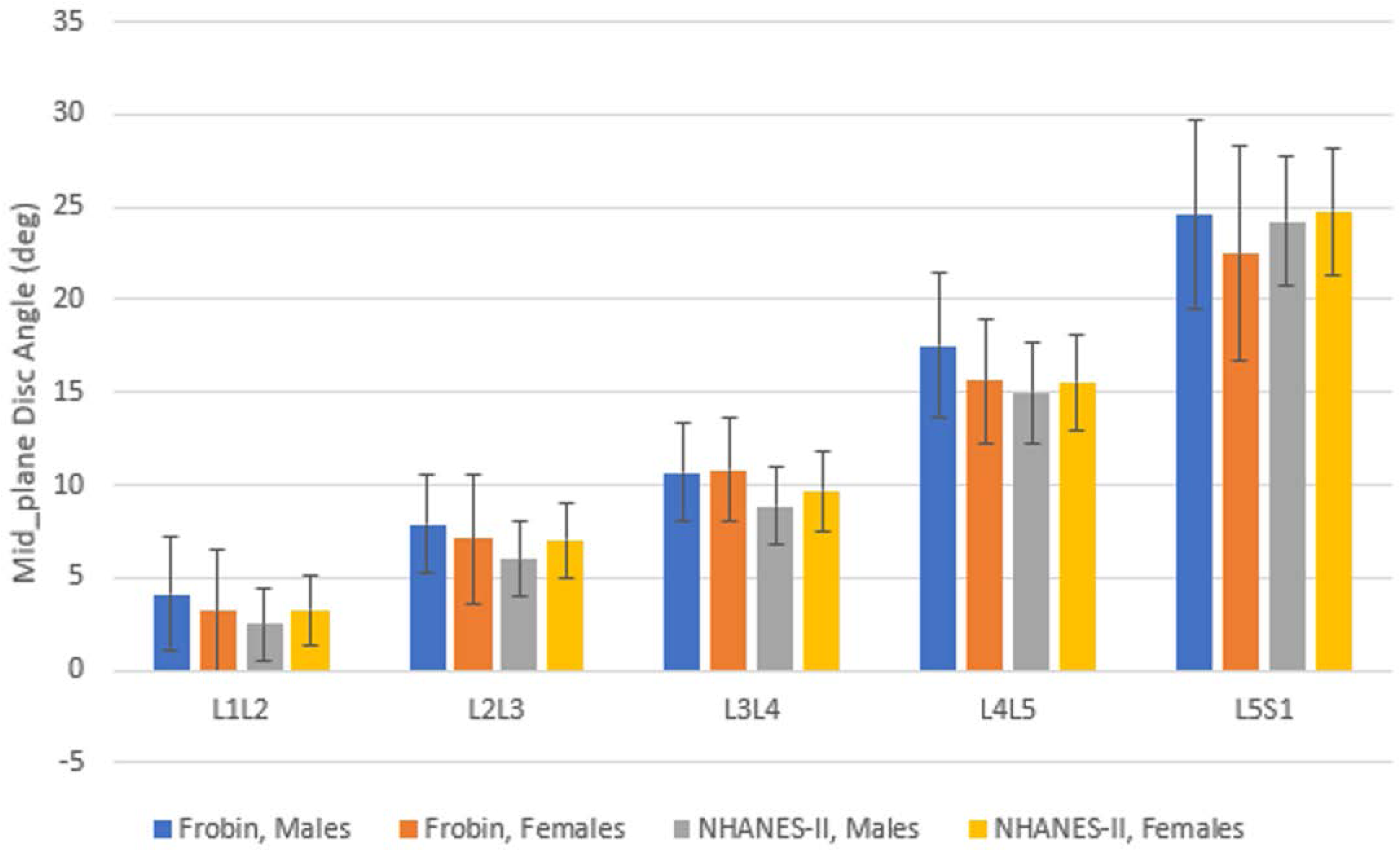
A comparison of mid-plane angles from the NHANES-II study (excluding all levels where any metric was abnormal) to data from Frobin et al.^14^ X-rays in the NHANES-II study were obtained with subjects lying on their sides in a semi flexed position, while Frobin et al obtained X-rays with patients standing. The error bars show standard deviations. The sample size in Frobin et al was 61 to 239 depending on sex and level, while the sample size in the NHANES-II data was 1185 to 2572 depending on sex and level.

**Table 4:** L1-S1 angles from three prior studies^70–72^ measured from standing lateral x-rays compared to L1-S1 angles in the NHANES-II study

|  | N | Mean Age | Mean L1-S1 | StdDev L1-S1 |
| --- | --- | --- | --- | --- |
| Korovessis<sup>70</sup> | 45 | 63 | 48 | 12.5 |
| Lord<sup>71</sup> | 109 | 47 | 49 | 15 |
| Been<sup>72</sup> | 100 | 38 | 51 |  |
| NHANES-II | 6397 | 55 | 51 | 13 |

From a practical standpoint, degenerative spondylolisthesis has traditionally been considered a single diagnosis. It is now clear that this in fact is a very heterogenous condition that requires a deeper understanding to effectively optimize its management. Beyond simply recognizing that this radiographic interpretation has a depth of complexity, we are also limited by our relatively primitive and inaccurate assessment tools. When a patient record or a scientific paper reports that a patient or patients had disc height loss or spondylolisthesis, what evidence exists that the report is accurate? What evidence exists that the magnitude of SPO is reliably reported? Will disc height and spondylolisthesis measurements prove valuable in diagnosis and treatment? It seems probable that clinical trials (past and future) might yield better and more actionable knowledge if accurate, reproducible, and standardized assessments are used. From a clinician’s perspective, there is something about spondylolisthesis that is contributing to clinical impairment in some but not all cases. Understanding the clinical predictors that forecast the role of spondylolisthesis in symptom generation both before and after surgery will be critical to optimal management of this prevalent condition. With developing artificial intelligence, it appears we are much closer to objectively and accurately evaluating and quantifying degenerative spondylolisthesis and automatically obtaining these metrics, with minimal effort, in routine practice. The NHANES-II reference data and measurements must be critically correlated with clinical metrics including but not limited to subjective complaints, physical exam, and sociodemographic features of the patient. Artificial intelligence will likely automate routine and basic measurements such as disc height loss and spondylolisthesis, such that the clinician can focus on integrating the assessments. Integrating imaging with clinical characteristics should allow development of reliable predictive analytics to optimize clinical and surgical decision making in the future.

Of note: there would be minimal technical challenges or cost to re-analysis of prior studies using objective SPO metrics to determine if the amount of SPO can help to predict treatment outcomes.

## Conclusion

The NHANES-II radiographs were used to develop reference data to aid in interpreting measurements of sagittal plane offset (SPO), disc heights, and disc angles. These metrics are dependent on disc level, so predicting normal SPO should always be done with level-specific reference data. The NHANES-II data document that SPO is significantly dependent on disc heights and disc angles, though the effect is small when all X-rays are obtained in a neutral position. Analysis of standing flexion-extension radiographs documents a far more significant effect of disc height and disc angle on SPO, and thus the importance of evaluating disc height and angle when predicting what should be normal SPO. The data support predicting normal SPO based only on level as long as there is little variability between subjects independent of the position of the spine, but disc heights and angles should be used to predict normal SPO when the spine is flexed or extended. The research and clinical value of any quantitative diagnostic is improved when sources of measurement error are minimized. Reference data developed from the NHANES-II lumbar X-rays provide normative reference data for predicting normal SPO, disc height, and disc angles. These reference data would need to be tested for efficacy in appropriate clinical trials. Multiple metrics were assessed using the NHANES-II data, yet it will take further research to determine which are clinically efficacious. The greatest challenge is to validate diagnosis and treatment algorithms that can effectively make use of disc and SPO metrics to improve outcomes for patients with spinal disorders.

## Ethics Approval

Vertebral morphology was measured from lateral lumbar and cervical radiographs obtained during the 2^nd^ National Health and Nutrition Survey (1976-1980). That study was designed and conducted to protect the rights of the volunteers, and all subjects in the study signed a consent as described in the study documentation:

https://wwwn.cdc.gov/nchs/nhanes/nhanes2/manualsandreports.aspx

https://wwwn.cdc.gov/nchs/data/nhanes2/manuals/15a76_79.pdf

## Data Availability

Sagittal plane offset, disc height, and disc angle data will be provided upon reasonable request.

## Acknowledgements

The publicly available NHANES-II database with images is gratefully acknowledged. Centers for Disease Control and Prevention (CDC). National Center for Health Statistics (NCHS). National Health and Nutrition Examination Survey Data. Hyattsville, MD: U.S. Department of Health and Human Services, Centers for Disease Control and Prevention. https://www.cdc.gov/nchs/data/nhanes/nhanes_release_policy.pdf<u>\</u>

## Dislosures

JAH, TFG, and PN are employees of Medical Metrics, Inc which is an imaging core laboratory with the technology used to make all of the radiographic measurements. They each received salary support to complete the work. No external funding was received in support of this work.

## Supplemental Appendix 1: Details of landmark placement, descriptive statistics, examples of abnormalities in the NHANES-II x-rays, and prevalence of abnormalities in the NHANES-II x-rays

### Details of vertebral landmark placement

The standardized landmark placement was focused on the mid-sagittal plane of each vertebral body (Figure 1), excluding any residual uncinate processes, and placing landmarks that identify the corners of the vertebral bodies prior to any osteophyte formation ^(1–3)^. The posterior superior landmarks are placed so as to represent the endplate as it would appear on a mid-sagittal slice of a CT exam, rather than up at the top of any posterior ridges on the upper endplate, similar to logic described in Keynan et al and Quint et al. ^(4,5)^.

When the X-ray beam path through a vertebra is not perpendicular to the mid-sagittal plane of the vertebra, the left and right rims of the vertebral endplates (as seen in Figure 1), and the left and right aspects of the posterior vertebral body may be seen on the X-ray.^(3,6)^ The mid-sagittal plane is assumed to be midway between the radiographic shadows of the left and right endplate rims, and midway between the radiographic shadows of the left and right aspects of the posterior wall. A similar approach was described by Quint et al.^(5)^ Multiple examples of landmark placement are provided online^1^ since disc space metrics calculated from landmarks are dependent on the nuances of how landmarks are placed, and it is therefore important to appreciate landmark placement details.

**Figure 1:**
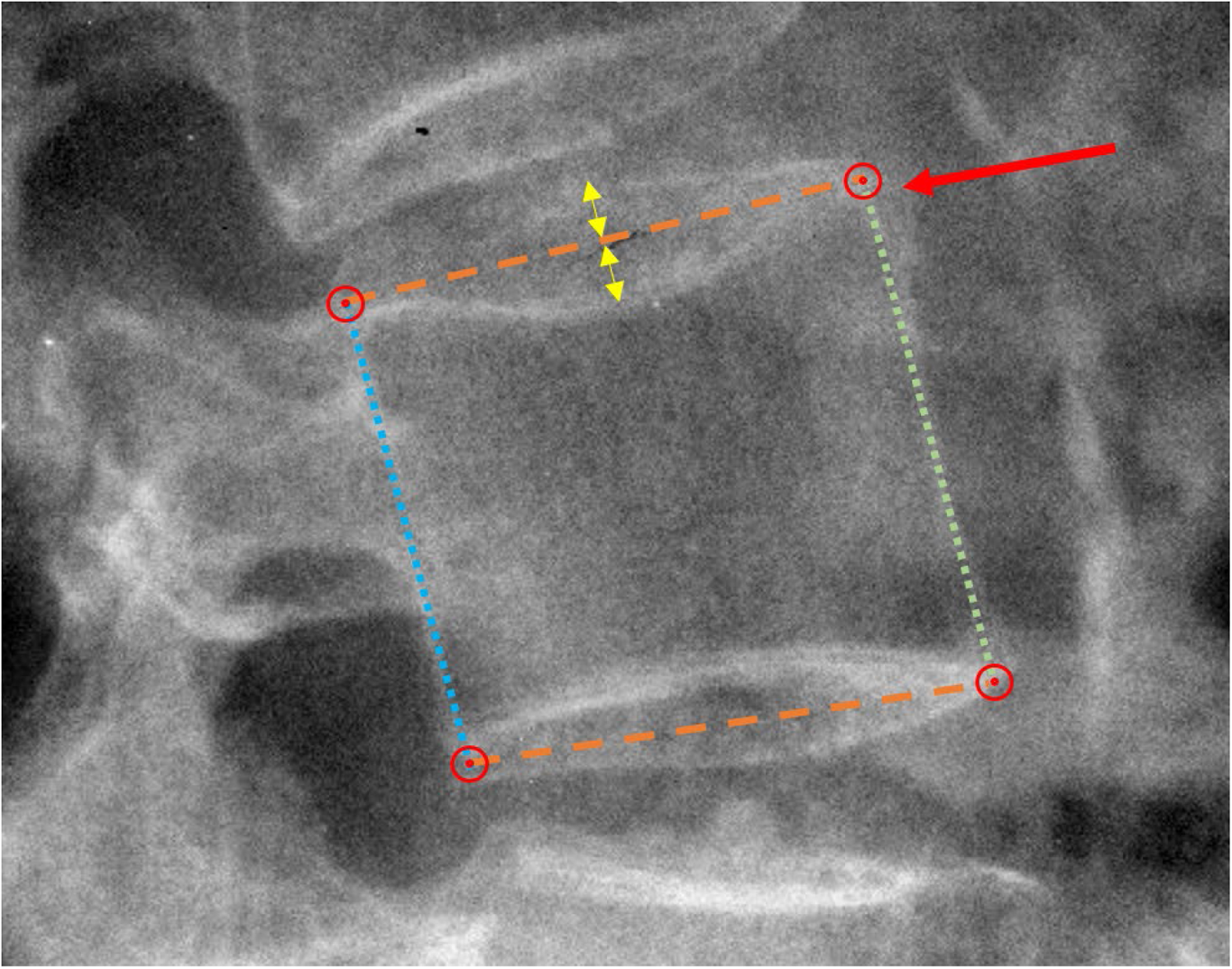
Details of anatomic landmark placement. Dashed lines show the estimated mid-sagittal plane of the superior and inferior endplates, identified as bisecting the radiographic shadows of the left and right sides of the endplates (yellow arrows). The red circles show the four landmarks used to measure vertebral body morphology and disc space. The red arrow points to an anterior osteophyte that is ignored when placing landmarks. The dotted green and blue lines show the anterior and posterior aspects of the vertebral body.

### Disc space and sagittal plane offset (SPO) metrics from the NHANES-II lumbar spine radiographs

**Table 1:** Descriptive statistics for Anterior disc heights (ADH), before and after [in brackets] trimming.

| Level | N | Mean | Std Dev | CV | Median | 10th | 95th | Skew | Kurtosis |
| --- | --- | --- | --- | --- | --- | --- | --- | --- | --- |
| L1L2 | 7020<br>[3487] | 25.47<br>[25.84] | 4.46<br>[2.75] | 0.17<br>[0.11] | 25.53<br>[25.82] | 20.04<br>[22.14] | 32.48<br>[30.44] | -0.28<br>[0.06] | 3.87<br>[2.31] |
| L2L3 | 7284<br>[3675] | 28.75<br>[29.36] | 4.89<br>[2.94] | 0.17<br>[0.10] | 29.07<br>[29.43] | 22.68<br>[25.29] | 36.16<br>[34.19] | -0.56<br>[-0.05] | 4.25<br>[2.32] |
| L3L4 | 7297<br>[3842] | 32.03<br>[32.71] | 5.31<br>[3.11] | 0.17<br>[0.09] | 32.36<br>[32.62] | 25.47<br>[28.59] | 39.90<br>[37.91] | -0.67<br>[-0.01] | 4.47<br>[2.42] |
| L4L5 | 7163<br>[3874] | 36.76[3<br>7.79] | 7.01<br>[3.99] | 0.19<br>[0.11] | 37.65<br>[37.95] | 27.54<br>[32.34] | 46.37<br>[44.07] | -0.76<br>[-0.22] | 4.07<br>[2.50] |
| L5S1 | 6682<br>[3573] | 41.38<br>[42.32] | 9.30<br>[5.84] | 0.22<br>[0.14] | 42.20<br>[42.57] | 28.41<br>[34.29] | 55.44<br>[51.56] | -0.36<br>[-0.18] | 2.96<br>[2.39] |

**Table 2:** Descriptive statistics for Posterior disc heights (PDH), before and after [in brackets] trimming.

| Level | N | Mean | Std Dev | CV | Median | 10th | 95th | Skew | Kurtosis |
| --- | --- | --- | --- | --- | --- | --- | --- | --- | --- |
| L1L2 | 7020<br>[3487] | 17.71<br>[17.97] | 3.17<br>[2.17] | 0.18<br>[0.12] | 17.67<br>[17.88] | 13.77<br>[15.03] | 22.97<br>[21.80] | 0.05<br>[0.15] | 3.57<br>[2.25] |
| L2L3 | 7284<br>[3675] | 17.12<br>[17.55] | 3.66<br>[2.44] | 0.21<br>[0.14] | 17.13<br>[17.47] | 12.46<br>[14.38] | 23.19<br>[21.72] | -0.03<br>[0.10] | 3.05<br>[2.27] |
| L3L4 | 7297<br>[3842] | 17.51<br>[18.01] | 3.91<br>[2.59] | 0.22<br>[0.14] | 17.58<br>[18.03] | 12.43<br>[14.48] | 23.73<br>[22.31] | -0.10<br>[-0.01] | 3.24<br>[2.23] |
| L4L5 | 7163<br>[3874] | 17.36<br>[18.01] | 4.55<br>[3.06] | 0.26<br>[0.17] | 17.57<br>[18.14] | 11.28<br>[13.75] | 24.56<br>[22.95] | -0.15<br>[-0.08] | 3.05<br>[2.26] |
| L5S1 | 6682<br>[3573] | 13.38<br>[13.83] | 4.73<br>[3.29] | 0.35<br>[0.24] | 13.22<br>[13.83] | 7.56<br>[9.39] | 21.33<br>[19.50] | 0.17<br>[0.13] | 3.30<br>[2.21] |

**Table 3:** Descriptive statistics for Average Disc Height (AvgDH), before and after [in brackets] trimming.

| Level | N | Mean | Std Dev | CV | Median | 10th | 90th | Skew | Kurtosis |
| --- | --- | --- | --- | --- | --- | --- | --- | --- | --- |
| L1L2 | 7020<br>[3436] | 21.59<br>[21.94] | 3.16<br>[2.08] | 0.15<br>[0.09] | 21.61<br>[21.83] | 17.70<br>[19.23] | 26.77<br>[25.45] | -0.23<br>[0.15] | 3.99<br>[2.35] |
| L2L3 | 7284<br>[3638] | 22.94<br>[23.49] | 3.63<br>[2.29] | 0.16<br>[0.10] | 23.18<br>[23.48] | 18.37<br>[20.37] | 28.55<br>[27.29] | -0.51<br>[0.04] | 3.95<br>[2.36] |
| L3L4 | 7297<br>[3811] | 24.77<br>[25.39] | 3.96<br>[2.47] | 0.16<br>[0.10] | 25.05<br>[25.36] | 19.75<br>[22.13] | 30.59<br>[29.41] | -0.65<br>[0.00] | 4.25<br>[2.35] |
| L4L5 | 7163<br>[3848] | 27.06<br>[27.94] | 5.00<br>[3.03] | 0.18<br>[0.11] | 27.71<br>[28.03] | 20.28<br>[23.86] | 33.97<br>[32.69] | -0.77<br>[-0.18] | 3.81<br>[2.46] |
| L5S1 | 6682<br>[3505] | 27.38<br>[28.16] | 5.74<br>[3.77] | 0.21<br>[0.13] | 28.01<br>[28.35] | 19.24<br>[22.93] | 35.96<br>[34.11] | -0.35<br>[-0.17] | 2.96<br>[2.49] |

**Table 4:** Descriptive statistics for Disc Angle (DA), before and after [in brackets] trimming.

| Level | N | Mean | Std Dev | CV | Median | 10th | 90th | Skew | Kurtosis |
| --- | --- | --- | --- | --- | --- | --- | --- | --- | --- |
| L1L2 | 7020<br>[3487] | 4.59<br>[4.64] | 2.63<br>[1.53] | 0.57<br>[0.33] | 4.61<br>[4.66] | 1.27<br>[2.57] | 7.90<br>[6.70] | -0.11<br>[-0.03] | 3.40<br>[2.25] |
| L2L3 | 7284<br>[3675] | 6.87<br>[6.96] | 2.78<br>[1.64] | 0.40<br>[0.24] | 6.87<br>[6.96] | 3.38<br>[4.70] | 10.35<br>[9.20] | -0.14<br>[-0.04] | 3.25<br>[2.20] |
| L3L4 | 7297<br>[3842] | 8.45<br>[8.54] | 2.88<br>[1.64] | 0.34<br>[0.19] | 8.52<br>[8.55] | 4.91<br>[6.33] | 12.04<br>[10.75] | -0.17<br>[-0.04] | 3.61<br>[2.26] |
| L4L5 | 7163<br>[3874] | 11.15<br>[11.36] | 3.66<br>[2.07] | 0.33<br>[0.18] | 11.37<br>[11.46] | 6.42<br>[8.52] | 15.51<br>[14.06] | -0.29<br>[-0.20] | 3.56<br>[2.31] |
| L5S1 | 6682<br>[3573] | 16.19<br>[16.49] | 5.37<br>[3.20] | 0.33<br>[0.19] | 16.41<br>[16.60] | 9.11<br>[12.08] | 23.04<br>[20.70] | -0.19<br>[-0.12] | 2.95<br>[2.19] |

**Table 5:** Descriptive statistics for Anterior Sagittal Plane Offset (ASPO), before and after [in brackets] trimming.

| Level | N | Mean | StdDev | CV | Median | 5th | 95th | Skew | Kurtosis |
| --- | --- | --- | --- | --- | --- | --- | --- | --- | --- |
| L1L2 | 7020<br>[3487] | -5.97<br>[-5.72] | 2.48<br>[1.64] | -0.42<br>[-0.29] | -5.81<br>[-5.65] | -10.31<br>[-8.63] | -2.29<br>[-3.10] | -0.44<br>[-0.27] | 4.44<br>[2.64] |
| L2L3 | 7284<br>[3675] | -7.40<br>[-7.22] | 2.86<br>[1.86] | -0.39<br>[-0.26] | -7.35<br>[-7.20] | -12.23<br>[-10.36] | -3.02<br>[-4.14] | 0.00<br>[-0.08] | 4.93<br>[2.50] |
| L3L4 | 7297<br>[3842] | -6.38<br>[-6.46] | 3.50<br>[2.10] | -0.55<br>[-0.33] | -6.47<br>[-6.47] | -11.65<br>[-9.95] | -0.75<br>[-2.85] | 0.70<br>[0.05] | 6.20<br>[2.66] |
| L4L5 | 7163<br>[3874] | -5.55<br>[-6.05] | 5.33<br>[3.05] | -0.96<br>[-0.50] | -6.32<br>[-6.34] | -12.37<br>[-10.55] | 4.01<br>[-0.16] | 1.52<br>[0.58] | 8.00<br>[3.18] |
| L5S1 | 6682<br>[3573] | -9.71<br>[-10.19] | 6.33<br>[3.70] | -0.65<br>[-0.36] | -10.31<br>[-10.43] | -18.50<br>[-15.90] | 0.89<br>[-3.40] | 1.31<br>[0.33] | 7.60<br>[2.80] |

**Table 6:** Descriptive statistics for Posterior Sagittal Plane Offset (PSPO), before and after [in brackets] trimming.

| Level | N | Mean | StdDev | CV | Median | 5th | 95th | Skew | Kurtosis |
| --- | --- | --- | --- | --- | --- | --- | --- | --- | --- |
| L1L2 | 7020<br>[3487] | 2.64<br>[2.55] | 2.32<br>[1.53] | 0.88<br>[0.60] | 2.56<br>[2.52] | -0.84<br>[0.05] | 6.46<br>[5.21] | 0.01<br>[0.15] | 5.60<br>[2.54] |
| L2L3 | 7284<br>[3675] | 3.97<br>[4.02] | 2.48<br>[1.66] | 0.62<br>[0.41] | 3.95<br>[4.01] | 0.08<br>[1.27] | 7.90<br>[6.88] | -0.29<br>[0.04] | 5.61<br>[2.45] |
| L3L4 | 7297<br>[3842] | 4.11<br>[4.34] | 3.06<br>[1.89] | 0.74<br>[0.44] | 4.31<br>[4.39] | -0.79<br>[1.13] | 8.49<br>[7.39] | -1.08<br>[-0.15] | 7.74<br>[2.50] |
| L4L5 | 7163<br>[3874] | 3.98<br>[4.53] | 4.75<br>[2.80] | 1.19<br>[0.62] | 4.74<br>[4.89] | -4.65<br>[-0.83] | 9.94<br>[8.56] | -1.86<br>[-0.59] | 10.38<br>[3.05] |
| L5S1 | 6682<br>[3573] | 5.99<br>[6.40] | 5.67<br>[3.28] | 0.95<br>[0.51] | 6.64<br>[6.56] | -3.24<br>[0.59] | 13.24<br>[11.52] | -1.93<br>[-0.34] | 11.27<br>[2.77] |

**Table 7:**
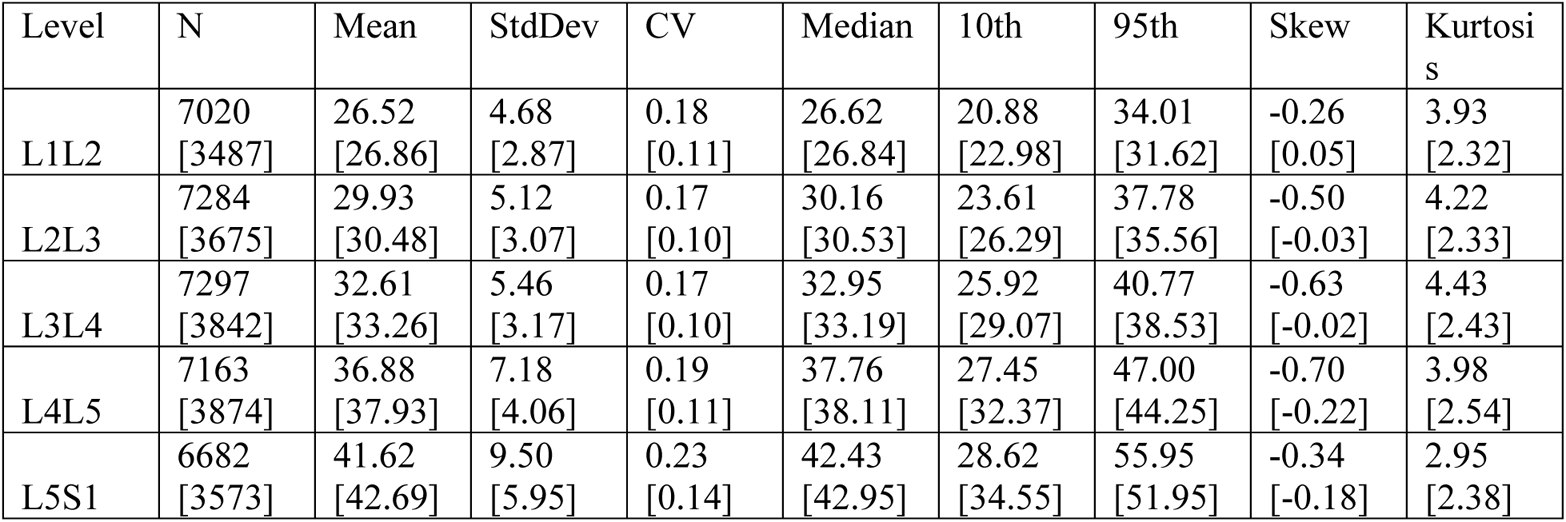
Descriptive statistics for Ventral disc heights (VDH), before and after [in brackets] trimming.

**Table 8:** Descriptive statistics for Dorsal disc heights (DDH), before and after [in brackets] trimming.

| Level | N | Mean | StdDev | CV | Median | 10th | 95th | Skew | Kurtosis |
| --- | --- | --- | --- | --- | --- | --- | --- | --- | --- |
| L1L2 | 7020<br>[3487] | 18.40<br>[18.63] | 3.25<br>[2.23] | 0.18<br>[0.12] | 18.35<br>[18.56] | 14.38<br>[15.64] | 23.80<br>[22.52] | 0.06<br>[0.14] | 3.63<br>[2.26] |
| L2L3 | 7284<br>[3675] | 17.81<br>[18.20] | 3.69<br>[2.46] | 0.21<br>[0.14] | 17.79<br>[18.13] | 13.12<br>[14.99] | 23.92<br>[22.38] | -0.03<br>[0.09] | 3.06<br>[2.27] |
| L3L4 | 7297<br>[3842] | 17.87<br>[18.36] | 3.87<br>[2.56] | 0.22<br>[0.14] | 17.94<br>[18.38] | 12.87<br>[14.84] | 23.98<br>[22.62] | -0.12<br>[-0.02] | 3.29<br>[2.23] |
| L4L5 | 7163<br>[3874] | 17.53<br>[18.20] | 4.53<br>[3.04] | 0.26<br>[0.17] | 17.78<br>[18.29] | 11.50<br>[13.99] | 24.62<br>[23.14] | -0.19<br>[-0.09] | 3.10<br>[2.27] |
| L5S1 | 6682<br>[3573] | 13.85<br>[14.34] | 4.53<br>[3.21] | 0.33<br>[0.22] | 13.74<br>[14.31] | 8.17<br>[10.02] | 21.48<br>[19.87] | 0.13<br>[0.10] | 3.19<br>[2.21] |

**Table 9:** Descriptive statistics for Mid-Plane Disc Angles (MPA), before and after [in brackets] trimming.

| Level | N | Mean | StdDev | CV | Median | 10th | 90th | Skew | Kurtosis |
| --- | --- | --- | --- | --- | --- | --- | --- | --- | --- |
| L1L2 | 7020<br>[3487] | 2.65<br>[2.74] | 3.27<br>[1.94] | 1.23<br>[0.71] | 2.70<br>[2.81] | -1.35<br>[0.09] | 6.75<br>[5.38] | -0.17<br>[-0.03] | 3.67<br>[2.20] |
| L2L3 | 7284<br>[3675] | 6.31<br>[6.34] | 3.41<br>[2.08] | 0.54<br>[0.33] | 6.32<br>[6.32] | 2.06<br>[3.50] | 10.56<br>[9.17] | -0.11<br>[-0.02] | 3.37<br>[2.13] |
| L3L4 | 7297<br>[3842] | 9.16<br>[9.13] | 3.59<br>[2.16] | 0.39<br>[0.24] | 9.15<br>[9.12] | 4.67<br>[6.15] | 13.62<br>[12.07] | 0.01<br>[0.03] | 3.43<br>[2.18] |
| L4L5 | 7163<br>[3874] | 15.16<br>[15.17] | 4.46<br>[2.67] | 0.29<br>[0.18] | 15.32<br>[15.25] | 9.55<br>[11.46] | 20.69<br>[18.67] | -0.11<br>[-0.06] | 3.51<br>[2.17] |
| L5S1 | 6682<br>[3573] | 24.07<br>[24.42] | 5.71<br>[3.49] | 0.24<br>[0.14] | 24.44<br>[24.65] | 16.44<br>[19.52] | 31.04<br>[29.01] | -0.27<br>[-0.19] | 3.11<br>[2.21] |

**Table 10:**
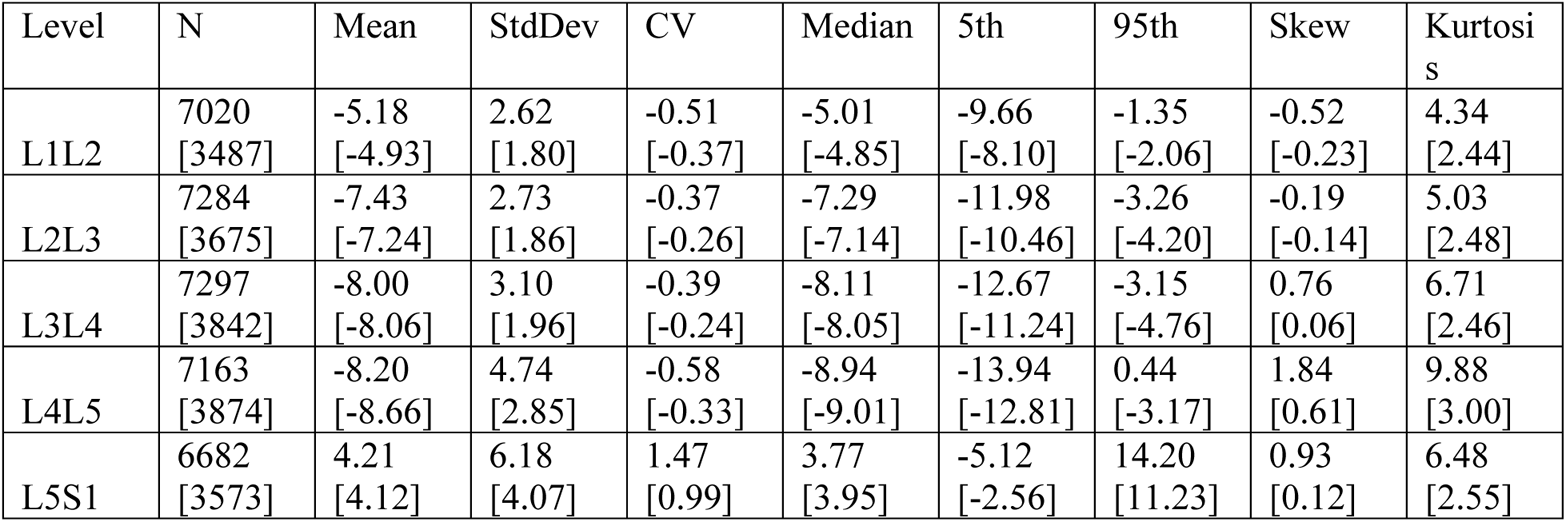
Descriptive statistics for Centroid Sagittal Plane Offset (CSPO), before and after [in brackets] trimming.

| Level | N | Mean | StdDev | CV | Median | 5th | 95th | Skew | Kurtosis |
| --- | --- | --- | --- | --- | --- | --- | --- | --- | --- |
| L1L2 | 7020<br>[3487] | -5.18<br>[-4.93] | 2.62<br>[1.80] | -0.51<br>[-0.37] | -5.01<br>[-4.85] | -9.66<br>[-8.10] | -1.35<br>[-2.06] | -0.52<br>[-0.23] | 4.34<br>[2.44] |
| L2L3 | 7284<br>[3675] | -7.43<br>[-7.24] | 2.73<br>[1.86] | -0.37<br>[-0.26] | -7.29<br>[-7.14] | -11.98<br>[-10.46] | -3.26<br>[-4.20] | -0.19<br>[-0.14] | 5.03<br>[2.48] |
| L3L4 | 7297<br>[3842] | -8.00<br>[-8.06] | 3.10<br>[1.96] | -0.39<br>[-0.24] | -8.11<br>[-8.05] | -12.67<br>[-11.24] | -3.15<br>[-4.76] | 0.76<br>[0.06] | 6.71<br>[2.46] |
| L4L5 | 7163<br>[3874] | -8.20<br>[-8.66] | 4.74<br>[2.85] | -0.58<br>[-0.33] | -8.94<br>[-9.01] | -13.94<br>[-12.81] | 0.44<br>[-3.17] | 1.84<br>[0.61] | 9.88<br>[3.00] |
| L5S1 | 6682<br>[3573] | 4.21<br>[4.12] | 6.18<br>[4.07] | 1.47<br>[0.99] | 3.77<br>[3.95] | -5.12<br>[-2.56] | 14.20<br>[11.23] | 0.93<br>[0.12] | 6.48<br>[2.55] |

### Proportion of levels with abnormalities. This includes all lumbar spines from the NHANES-II study. Abnormalities were identified by standardized (Z) scores in units of std dev from average normal

**Table 11:** Proportion with disc heights > normal

| Level | zADH>2 | zADH>3 | zPDH>2 | zPDH>3 | zVDH>2 | zVDH>3 | zDDH>2 | zDDH>3 |
| --- | --- | --- | --- | --- | --- | --- | --- | --- |
| L1L2 | 8.4 | 2.4 | 7.2 | 1.7 | 8.7 | 2.5 | 7.6 | 1.8 |
| L2L3 | 7.4 | 1.7 | 7.5 | 1.7 | 7.9 | 2.1 | 7.6 | 1.8 |
| L3L4 | 7.4 | 1.7 | 6.6 | 1.7 | 8.2 | 2.0 | 6.4 | 1.7 |
| L4L5 | 6.5 | 1.4 | 6.1 | 1.2 | 7.0 | 1.7 | 5.9 | 1.1 |
| L5S1 | 7.0 | 1.2 | 7.3 | 1.8 | 7.0 | 1.2 | 6.5 | 1.5 |

**Table 12:** Proportions with disc Heights < normal

| Level | zADH<-2 | zADH<-3 | zPDH<-2 | zPDH<-3 | zVDH<-2 | zVDH<-3 | zDDH<-2 | zDDH<-3 |
| --- | --- | --- | --- | --- | --- | --- | --- | --- |
| L1L2 | 11.2 | 4.1 | 9.1 | 2.3 | 11.0 | 4.0 | 8.8 | 2.3 |
| L2L3 | 12.4 | 5.2 | 11.1 | 3.2 | 12.0 | 5.0 | 10.6 | 3.2 |
| L3L4 | 13.5 | 5.9 | 11.6 | 3.6 | 13.1 | 5.9 | 11.5 | 3.6 |
| L4L5 | 14.4 | 7.6 | 12.0 | 3.6 | 14.5 | 7.6 | 12.4 | 3.8 |
| L5S1 | 13.9 | 5.0 | 8.9 | 1.8 | 14.1 | 5.1 | 9.0 | 1.8 |

**Table 13:** Proportions with disc angles > normal

| Level | zDA>2 | zDA>3 | zMPA>2 | zMPA>3 |
| --- | --- | --- | --- | --- |
| L1L2 | 11.4 | 3.8 | 10.7 | 3.1 |
| L2L3 | 10.8 | 3.2 | 10.2 | 3.2 |
| L3L4 | 11.3 | 3.8 | 11.0 | 3.9 |
| L4L5 | 10.0 | 2.7 | 10.6 | 3.1 |
| L5S1 | 10.4 | 2.7 | 8.8 | 2.0 |

**Table 14:** Proportions with disc angles < normal

| Level | zDA<-2 | zDA<-3 | zMPA<-2 | zMPA<-3 |
| --- | --- | --- | --- | --- |
| L1L2 | 12.2 | 4.3 | 11.2 | 3.9 |
| L2L3 | 11.9 | 4.3 | 10.8 | 3.5 |
| L3L4 | 12.2 | 4.8 | 10.5 | 3.3 |
| L4L5 | 13.9 | 5.9 | 11.2 | 4.1 |
| L5S1 | 13.4 | 4.8 | 12.8 | 4.8 |

**Table 15:** Proportions with SPO > normal

| Level | zASPO>2 | zASPO>3 | zPSPO>2 | zPSPO>3 | zCSPO>2 | zCSPO>3 |
| --- | --- | --- | --- | --- | --- | --- |
| L1L2 | 5.6 | 1.1 | 9.3 | 3.0 | 4.9 | 0.8 |
| L2L3 | 7.0 | 1.9 | 7.7 | 2.1 | 6.1 | 1.4 |
| L3L4 | 8.9 | 4.0 | 6.3 | 1.7 | 7.9 | 3.5 |
| L4L5 | 11.3 | 5.8 | 4.3 | 0.6 | 9.9 | 5.5 |
| L5S1 | 10.9 | 5.0 | 5.7 | 1.0 | 7.7 | 3.6 |

**Table 16:** Proportions with SPO < normal

| Level | zASPO<-2 | zASPO<-3 | zPSPO<-2 | zPSPO<-3 | zCSPO<-2 | zCSPO<-3 |
| --- | --- | --- | --- | --- | --- | --- |
| L1L2 | 10.6 | 4.0 | 6.4 | 1.6 | 9.6 | 3.4 |
| L2L3 | 9.8 | 3.5 | 7.5 | 2.3 | 9.2 | 3.0 |
| L3L4 | 8.7 | 2.9 | 9.1 | 4.0 | 7.4 | 2.0 |
| L4L5 | 5.6 | 1.0 | 10.7 | 5.6 | 3.8 | 0.6 |
| L5S1 | 7.2 | 1.2 | 9.3 | 4.7 | 6.8 | 1.6 |

## Supplemental Appendix 2: Measurement of spondylolisthesis in flexion and extension

### Background and Methods

Although disc heights and disc angles significantly explained some of the variance in Sagittal Plane Offset (SPO) metrics in the NHANES-II data, the influence was relatively small (best R^2^ for anterior SPO was 0.56) and much of the variability in SPO was left unexplained.. The NHANES-II radiographs were obtained with subjects side-lying with relatively little variation in L1-S1 angle expected between subjects, compared to what can be expected in flexion-extension radiographs obtained with good subject effort. Spondylolisthesis may sometimes be assessed from flexion and extension (FE) radiographs in clinical practice and research studies^1^. Therefore, Anterior SPO (ASPO), Posterior SPO (PSPO) and Centroid SPO (CSPO) was also analyzed in flexion-extension radiographs of asymptomatic volunteers.

Some (162) of the FE radiographs were described and analyzed in a prior publication.^2^ An additional 211 FE radiographs were also analyzed. The first 162 FE studies had been obtained using a seated protocol ^2^, while the second 211 had been obtained with an upright protocol using a standard walker for support. Each of the 211 subjects watched a short training video before the X-rays were collected^1^. All 373 subjects consented to the IRB approved studies, were asymptomatic, and had never had medical treatment for a back disorder.

The FE studies were analyzed using the same series of neural networks and coded logic (Spine CAMP^TM^, Medical Metrics, Inc., Houston, TX) as with the NHANES-II X-rays. In addition, three highly experienced musculoskeletal radiologists assessed disc degeneration at every level using the Kellgren-Lawrence (KL) grading system.^3,4^ The first two radiologists assessed all radiographs, and the third adjudicated if the first two did not agree on the KL grade. The nine SPO and disc metrics were calculated from the landmarks using only those levels with no more than KL grade 1 (doubtful) disc degeneration. Regression analysis was used to obtain an equation to predict ASPO from level, ADH, and PDH. (Stata ver 15, College Station, TX).

### Results

Applying the normative data from the NHANES-II study (where all subjects were imaged in a side-lying position) to the 373 flexion-extension radiographs, 45% of the flexion and extension radiographs have standardized ASPO metrics >2 or < −2 Std Dev from average normal ASPO. Since 45% of asymptomatic levels can’t have abnormal SPO, this documents that normative SPO reference data established using the NHANES-II data should NOT be used with flexion and extension radiographs, at least with a threshold of >2 or < −2 Std Dev from average normal ASPO to classify SPO as abnormal.

To establish a method for measuring SPO in the presence of flexion or extension, the disc height, disc angle, and SPO data for the flexion and extension radiographs were trimmed to avoid including abnormalities. Trimming of the flexion-extension data was based on both SPO metrics (<5^th^ or > 95^th^ percentiles excluded) and the radiologist Kellgren-Lawrence (KL) grades to identify abnormal or degenerated levels. All levels in the flexion-extension data were trimmed where ASPO, PSPO, or CSPO was abnormal or where radiographic disc degeneration was > “doubtful” (KL grade 1). Based on the radiologist’s grading of disc degeneration combined with the SPO metrics, 25% of levels had degeneration but normal SPO, 11.3% had abnormal SPO but no degeneration, and 7.8% had both degeneration and abnormal SPO. Thus, 44.2% of levels were excluded from establishing an equation to predict normal SPO from disc heights and disc angles.

Analysis of variance revealed that the combination of ADH, PDH, and DA predicted 72% of the variation in PSPO and 85% of the variation in ASPO. The combination of VDH, DDH and MPA predicted 71% of the variation in CSPO. Since the R^2^ for ASPO was the highest, a regression equation was developed for predicting normal ASPO from ADH, PDH, and DA. Using the trimmed data, the Equation 1:

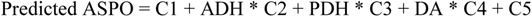

Where:

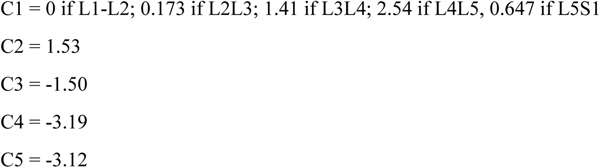

As a check of the regression equation, the predicted ASPO was linearly (P<0.0001) correlated to the actual ASPO for the trimmed data (R^2^ = 0.85). The average difference between the predicted ASPO and the actual ASPO, using only the trimmed NHANES-II data, was 0.014 % endplate width (std dev 2.1). The differences ranged from −6.3 to 11.4 % endplate width. Including all of the NHANES-II data, the difference between the predicted (from Equation 1) and actual ASPO was strongly (R^2^ = 0.64) and linearly (P<0.0001) correlated to the standardized ASPO metric (Figure 1). A stronger correlation would not be expected, since the predicted ASPO is the predicted **normal** ASPO, and actual ASPO in the NHANES-II study is not always normal. It is the difference between the actual and predicted (normal) ASPO that could be of clinical value. This supports the potential value of the ASPO prediction equation from the asymptomatic flexion-extension X-Rays. Further research will be required to determine which metric is best for classifying SPO as normal versus abnormal: the standardized ASPO, PSPO, or CSPO metric from the NHANES-II study, or the prediction equation derived from the flexion-extension studies. Nevertheless, this supports the hypothesis that it is necessary to account for disc heights and disc angle when predicting what normal SPO should be, particularly if there is a lot of flexion or extension.

**Figure 1:**
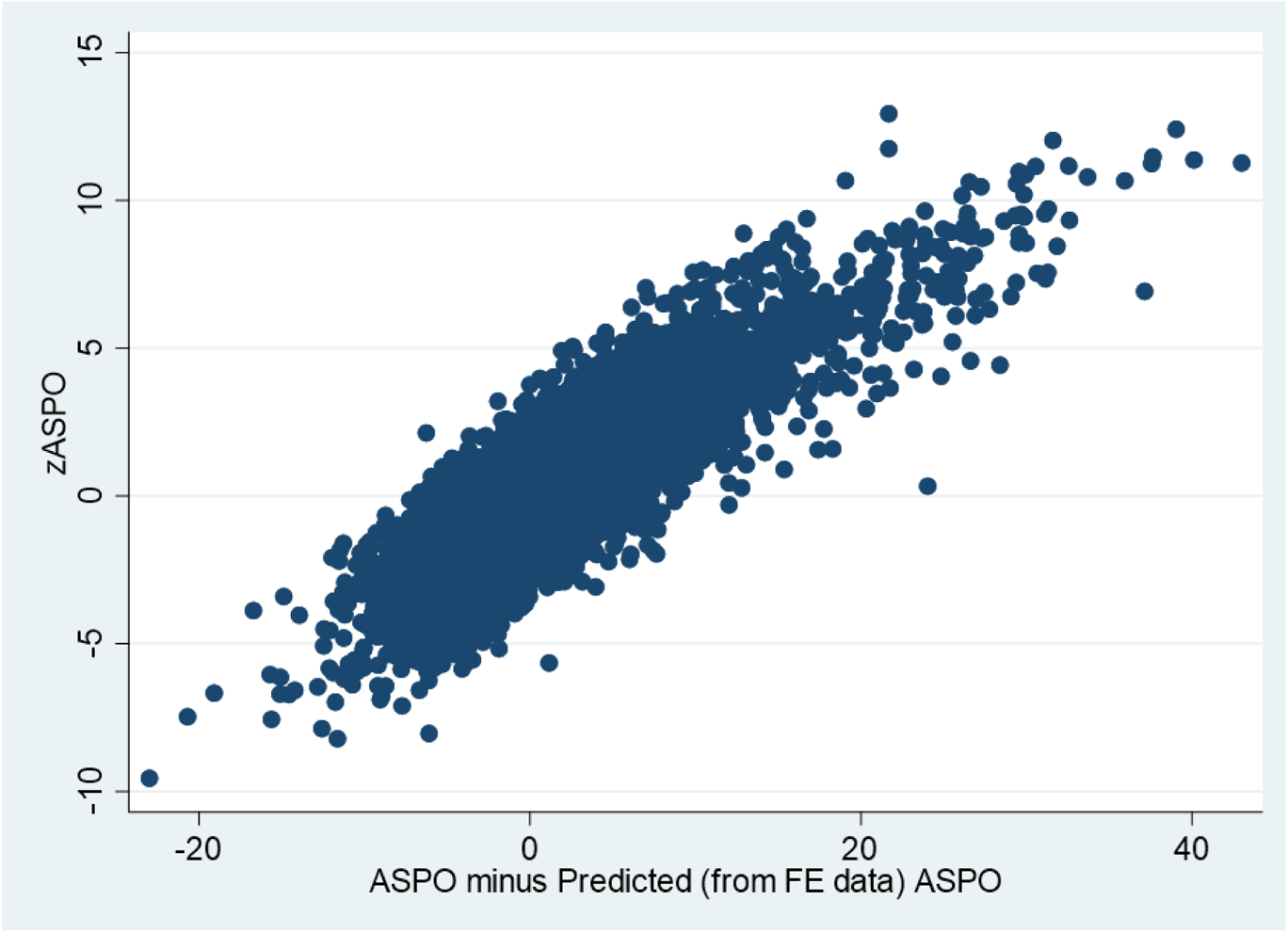
On the X-axis is the difference (units of precent endplate width) between the actual ASPO and the predicted ASPO (using the prediction equation derived from flexion-extension radiographs). On the Y-axis is the standardized ASPO (standard deviations from average ASPO in the trimmed NHANES-II data).

ASPO predicted using the flexion-extension X-rays can also be expressed as a standardized metric (difference from average normal). Since the predicted ASPO is calculated from an equation, a different approach to standardization is used. To obtain the denominator needed to calculate a standardized predicted ASPO, the standard error of the forecast was calculated after the regression. The standard error of the forecast is the standard error of the point prediction for 1 observation. There was minimal difference between levels, so one average value for all levels was used (1.87). A standardized SPO that accounts for disc heights and angles was then calculated as the actual ASPO minus the predicted ASPO divided by the standard error of the forecast. This is referred to as the spondylolisthesis index (SI) to distinguish it from the other SPO metrics described in the NHANES-II study.

To assess the difference in SPO between flexion and extension, the SI was calculated for every level for both the flexion and extension x-rays. Table 1 provides the median, 5^th^ and 95^th^ percentile differences between standardized (using NHANES-II data) ASPO, PSPO, and CSPO calculated in flexion versus extension. The difference in SI in flexion versus extension is also provided. Very large differences were observed between flexion and extension in the NHANES-II standardized ASPO, PSPO, and CSPO metrics. This also supports that the SPO metrics from the NHANES-II study should NOT be used for flexion-extension radiographs. Smaller differences were observed in the SI. Note that there were some cases in the flexion-extension X-rays of asymptomatic volunteers with clear spondylolisthesis. The data document that the median SI changes by approximately 1 between flexion and extension in an asymptomatic population, and that there are some levels with minimal change and others with substantial change. Further research is needed to determine if this can be of clinical value.

**Table 1:**
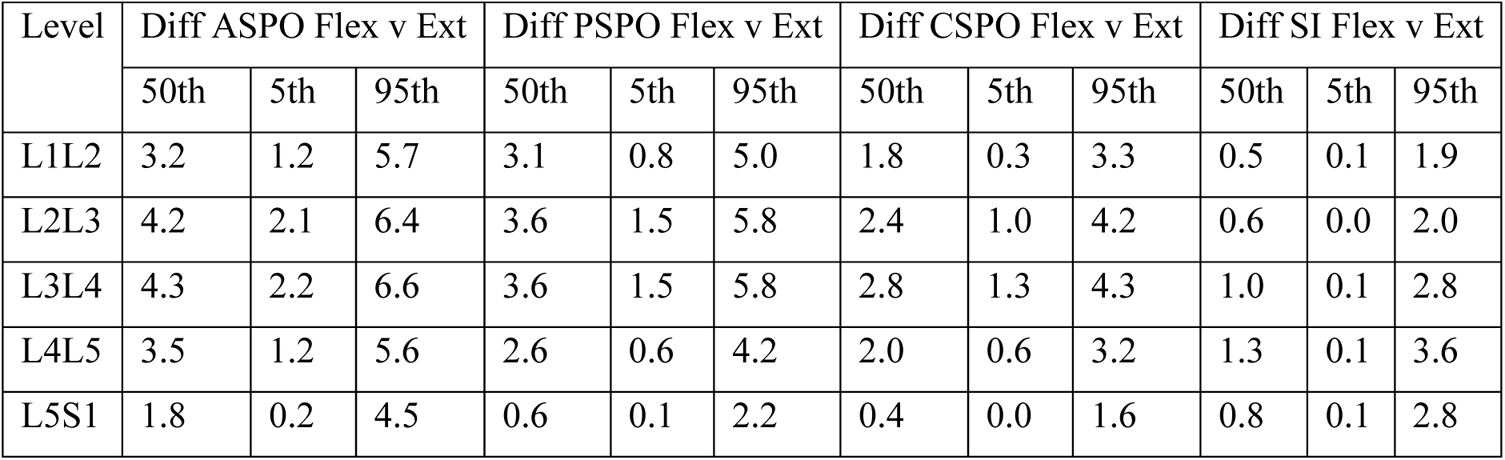
Differences in the <u>standardized</u> SPO metrics when the metric in flexion is compared to the same metric in extension. Note the large variations in ASPO, PSPO, and CSPO compared to the SI.

## Supplemental Appendix 3: Analysis of errors due to out-of-plane imaging

### Materials and Methods

To document expected variability in the metrics due to variations in radiographic projections that could occur in clinical practice, digitally reconstructed radiographs (DRR) were used. Thin-slice (1 mm or less), anonymized computed tomography exams of the lumbar spine of 26 individuals were interpolated to 0.2 mm isotropic resolution. The interpolation was done to achieve higher resolution DRRs. The three-dimensional (3D) coordinates of the four anatomic landmarks described in Supplementary Appendix 1, Figure 1 were manually digitized in the mid-sagittal plane of each vertebra from L1 to S1. Landmark position was identified using intersecting axial, coronal, and sagittal slices. Landmarks were thus placed in the mid-sagittal plane at clear and well-defined locations. The image processing and landmark placement were completed using Slicer 3D.^1^ The nine disc and SPO metrics described in the paper were calculated directly from the 3D landmark coordinates and used as the “gold standard” to assess accuracy of measurements from the DRRs.

Custom python code was developed to create 2D, lateral, digitally reconstructed radiographs from the 3D CT data using the Plastimatch DRR function^2^. A 40 inch source-to-image distance was used for all X-rays. The base x-ray was centered on the centroid of the L3 vertebra (this is the baseline isocenter). The distance between the center of L3 and the image was set to ½ the estimated width of the hips. The projection matrices used by Plastimatch DRR to create the simulated X-rays were then used to calculate, from the known 3D coordinates, the precise coordinates of each landmark on each 2D simulated X-ray. In addition to the baseline DRR, forty additional DRRs were generated with the following variations:

- random beam tilts of between ± 10 deg in each of the sagittal and axial planes
- random displacements from the baseline isocenter of ± 2 endplate widths in the AP and LR directions, plus ± 3 endplate width shifts in the cranial-caudal direction
- random image rotations between ± 24 deg

These variations resulted in a wide range of radiographic projections, and represent a digital version of the methodology used in earlier studies that addressed the effect of radiographic projection on disc height measurements. ^3–5^ The landmarks in every image were precisely calculated so all 2D landmark coordinates corresponded to the true 3D coordinates. This allows for documenting the effect of errors in vertebral morphology metrics that are due to radiographic projection, without uncertainty in landmark placement. These precisely calculated landmarks were analyzed to obtain the nine disc height, disc angle, and SPO metrics described in the paper. There were thus 41 measurements for every disc level (L1-L2 to L5-S1) for each of the 26 independent CT exams.

Although the coordinates of landmarks in every simulated X-ray were precisely calculated based on geometry, the quality of the simulated radiographs was below clinical standards, did not include the field-of-view normally found in a lateral spine X-ray, and were therefore not appropriate for use toward validating the accuracy of neural networks in obtaining landmarks. It should also be appreciated that the simulated radiographs with no beam tilt or offset cannot be assumed to be the optimal radiograph, since not all CT exams were obtained with perfectly oriented spines and some spines had frontal plane curvatures in the lumbar spine.

### Results

Table 1 provides the median, 5^th^ and 95^th^ percentile errors in the metrics. These are errors solely due to variability in radiographic projection with no error in landmark digitization. Standardized metrics (units of Std Dev from average normal) are reported in Table 1 to facilitate interpretation of the data and allow for pooling of data for all levels. For example, the median error in ASPO, due to random variations in radiographic projection, was 0.19 std dev from average ASPO in healthy discs. The maximum error in ASPO that occurred due to variations in radiographic projection was 1.1 std dev.

### Discussion

In general, errors due to variability in radiographic projection are small. However, large errors can occur with severe out-of-plane imaging. Disc angles were the least effected, followed by disc heights and then SPO. The worst errors were found in CT exams of scoliotic spines (eg Figure 1). With the spine in Figure 1, optimal imaging of the L2L3 level would require substantial beam tilt, and that amount of beam tilt would yield poor quality imaging of the L4L5 level.

### Conclusion

This experiment documents the need for caution when interpreting sagittal plane disc and SPO metrics when the radiographic projection is substantially out-of-plane. The images can be out-of-plane both with respect to the frontal and axial planes. A method to determine when there is too much out-of-plane is challenging. This requires both a definition of how much error is too much, and a method to detect out-of-plane that would result in too much error. Neural networks can likely be trained to recognize excessive out-of-plane once error tolerances are defined.

**Table 1:**
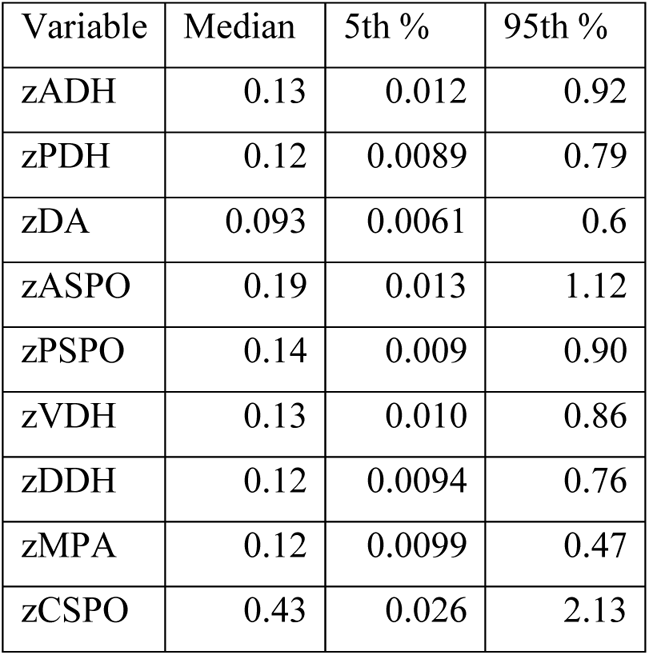
Descriptive statistics to help assess the effect of radiographic projection on error in the metrics. The absolute error for each metric was measured by comparing metrics from 2D simulated radiographs to the known measurement made from 3D data. Errors were analyzed using standardized (“z”) scores in units of std devs from average in healthy discs. Over 5,000 measurements were available for each metric (26 CT exams * 5 levels * 41 variations in radiographic projection).

**Figure 1:**
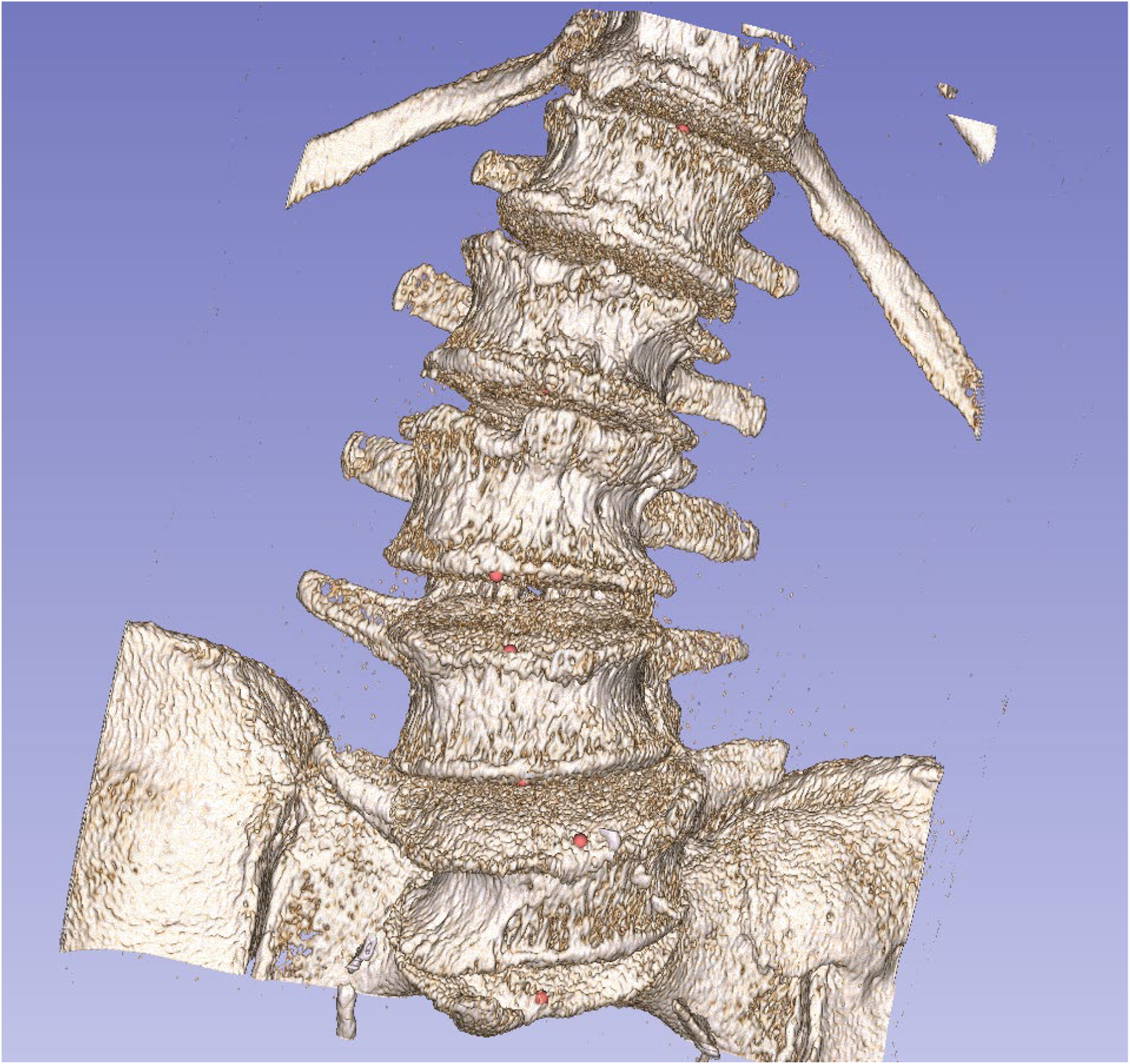
Three-dimensional reconstruction of one of the spines in the simulated X-ray study. Considering how a lateral radiograph of the lumbar spine would appear, helps to understand how disc heights and angles could change as the x-ray beam angle changes in the frontal plane.

## Footnotes

1 http://www.wheelessonline.com/ISSLS/section-15-chapter-2-radiology-and-degenerative-spondylolisthesis/

2 https://www.cdc.gov/nchs/nhanes/index.htm

1 https://www.dropbox.com/sh/qzrocrh86goxarx/AAADRXs5HoEjVdRcGIwA4beIa?dl=0

1 https://youtu.be/KaEQP57qWgU <u>English language version</u> https://youtu.be/Day_wvEG-yI <u>Spanish language version</u>

## Notes

### Competing Interest Statement

Several of the authors are employees of Medical Metrics, Inc which is an imaging core laboratory with the technology used to make all of the radiographic measurements. They each received salary support to complete the work. No external funding was received in support of this work.

### Funding Statement

No funding was received in support of this research other than salary support for some of the authors.

### Author Declarations

This study used the publicly available NHANES-II database with images: Centers for Disease Control and Prevention (CDC). National Center for Health Statistics (NCHS). National Health and Nutrition Examination Survey Data. Hyattsville, MD: U.S. Department of Health and Human Services, Centers for Disease Control and Prevention. https://www.cdc.gov/nchs/data/nhanes/nhanes_release_policy.pdf\

## References

1. Galbusera F, Casaroli G, Bassani T. Artificial intelligence and machine learning in spine research. Jor Spine. 2019;2(1):e1044.

2. Chang M, Canseco JA, Nicholson KJ, Patel N, Vaccaro AR. The role of machine learning in spine surgery: the future is now. Frontiers in surgery. 2020;7:54.

3. Ren G, Yu K, Xie Z, et al. Current Applications of Machine Learning in Spine: From Clinical View. Global Spine Journal. 2021:21925682211035363.

4. Câmara JR, Keen JR, Asgarzadie F. Functional radiography in examination of spondylolisthesis. American Journal of Roentgenology. 2015;204(4):W461–W469.

5. McDowell A. Plan and operation of the second National Health and Nutrition Examination Survey, 1976-1980. 1981;

6. Zhao KD, Ben-Abraham EI, Magnuson DJ, et al. Effect of Off-Axis Fluoroscopy Imaging on Two-Dimensional Kinematics in the Lumbar Spine: A Dynamic In Vitro Validation Study. J Biomech Eng. May 2016;138(5):054502. doi:10.1115/1.4032995

7. Zhao KD, Yang C, Zhao C, Stans AA, An KN. Assessment of noninvasive intervertebral motion measurements in the lumbar spine. JBiomechanics. 2005;38(9):1943–1946. NOT IN FILE. doi:10.1016/j.jbiomech.2004.07.029

8. Reitman CA, Hipp JA, Nguyen L, Esses SI. Changes in segmental intervertebral motion adjacent to cervical arthrodesis: a prospective study. Spine (Phila Pa 1976). Jun 1 2004;29(11):E221–6. doi:00007632-200406010-00022 [pii]

9. Reitman CA, Mauro KM, Nguyen L, Ziegler JM, Hipp JA. Intervertebral motion between flexion and extension in asymptomatic individuals. Spine (Phila Pa 1976). Dec 15 2004;29(24):2832–43. doi:00007632-200412150-00009 [pii]

10. Patwardhan AG, Havey RM, Wharton ND, et al. Asymmetric motion distribution between components of a mobile-core lumbar disc prosthesis: an explanation of unequal wear distribution in explanted CHARITE polyethylene cores. The Journal of bone and joint surgery American volume. May 2 2012;94(9):846–54. doi:10.2106/JBJS.J.00638

11. Pearson AM, Spratt KF, Genuario J, et al. Precision of lumbar intervertebral measurements: Does a computer-assisted technique improve reliability? Spine. 2011;36(7):572–580. doi:10.1097/BRS.0b013e3181e11c13

12. Pfeiffer M, Geisel T. Analysis of a computer-assisted technique for measuring the lumbar spine on radiographs: correlation of two methods. Academic radiology. 2003;10(3):275–282.

13. Hipp JA, Grieco TF, Newman P, Reitman CA. Definition of normal vertebral morphology using NHANES-II radiographs. medRxiv. 2022;

14. Frobin W, Brinckmann P, Biggemann M, Tillotson M, Burton K. Precision measurement of disc height, vertebral height and sagittal plane displacement from lateral radiographic views of the lumbar spine. Clin Biomech (Bristol, Avon). 1997;12 Suppl 1:S1–S63. doi:10.1016/s0268-0033(96)00067-8

15. Frobin W, Brinckmann P, Leivseth G, Biggemann M, Reikeras O. Precision measurement of segmental motion from flexion-extension radiographs of the lumbar spine. ClinBiomech(Bristol, Avon). 1996;11(8):457–465. NOT IN FILE.

16. Ravi B, Rampersaud R. Clinical magnification error in lateral spinal digital radiographs. Spine. 2008;33(10):E311–E316.

17. Shigematsu H, Koizumi M, Yoneda M, Iida J, Oshima T, Tanaka Y. Magnification error in digital radiographs of the cervical spine against magnetic resonance imaging measurements. Asian spine journal. 2013;7(4):267.

18. Lunt M, Gowin W, Johnell, Armbrecht G, Felsenberg D. A statistical method to minimize magnification errors in serial vertebral radiographs. OsteoporosInt. 2001;12(11):909–913. NOT IN FILE.

19. Staub BN, Holman PJ, Reitman CA, Hipp JA. Sagittal plane lumbar intervertebral motion during seated flexion-extension in 658 asymptomatic, non-degenerated levels. J Neurosurgery Spine. 2015 2015;23(6):731–738. doi:10.3171/2015.3.SPINE14898

20. Bifulco P, Sansone M, Cesarelli M, Allen R, Bracale M. Estimation of out-of-plane vertebra rotations on radiographic projections using CT data: a simulation study. MedEng Phys. 2002;24(4):295–300. NOT IN FILE.

21. Andersson G, Schultz A, Nathan A, Irstam L. Roentgenographic measurement of lumbar intervertebral disc height. Spine. 1981;6(2):154–158.

22. Danielson B, Frennered K, Irstam L. Roentgenologic assessment of spondylolisthesis: I. A study of measurement variations. Acta Radiologica. 1988;29(3):345–351.

23. McCarty ME, Mehlman CT, Tamai J, Do TT, Crawford AH, Klein G. Spondylolisthesis: intraobserver and interobserver reliability with regard to the measurement of slip percentage. Journal of Pediatric Orthopaedics. 2009;29(7):755–759.

24. Watters WC, 3rd, Bono CM, Gilbert TJ, et al. An evidence-based clinical guideline for the diagnosis and treatment of degenerative lumbar spondylolisthesis. Spine J. Jul 2009;9(7):609–14. doi:10.1016/j.spinee.2009.03.016

25. Matz PG, Meagher R, Lamer T, et al. Guideline summary review: an evidence-based clinical guideline for the diagnosis and treatment of degenerative lumbar spondylolisthesis. The Spine Journal. 2016;16(3):439–448.

26. Mummaneni PV, Bisson EF, Kerezoudis P, et al. Minimally invasive versus open fusion for Grade I degenerative lumbar spondylolisthesis: analysis of the Quality Outcomes Database. Neurosurgical focus. 2017;43(2):E11.

27. Bourassa-Moreau É, Mac-Thiong J-M, Labelle H. Redefining the technique for the radiologic measurement of slip in spondylolisthesis. Spine. 2010;35(14):1401–1405.

28. Danielson B, Frennered K, Selvik G, Irstam L. Roentgenologic Assessment of Spondylolisthesis: II. An Evaluation of Progression. Acta Radiologica. 1989;30(1):65–68.

29. Wall MS, Oppenheim WL. Measurement error of spondylolisthesis as a function of radiographic beam angle. Journal of pediatric orthopedics. 1995;15(2):193–198.

30. Tallroth K, Ylikoski M, Landtman M, Santavirta S. Reliability of radiographical measurements of spondylolisthesis and extension-flexion radiographs of the lumbar spine. Eur J Radiol. 1994;18(3):227–231. NOT IN FILE.

31. Mac-Thiong JM, Duong L, Parent S, et al. Reliability of the Spinal Deformity Study Group classification of lumbosacral spondylolisthesis. Spine (Phila Pa 1976). Jan 15 2012;37(2):E95–102. doi:10.1097/BRS.0b013e3182233969

32. Bao H, Yan P, Zhu W, et al. Validation and reliability analysis of the spinal deformity study group classification for L5-S1 lumbar spondylolisthesis. Spine. 2015;40(21):E1150–E1154.

33. Denard PJ, Holton KF, Miller J, et al. Lumbar spondylolisthesis among elderly men: prevalence, correlates and progression. Spine. 2010;35(10):1072.

34. Bolesta MJ, Winslow L, Gill K. A comparison of film and computer workstation measurements of degenerative spondylolisthesis: intraobserver and interobserver reliability. Spine. 2010;35(13):1300–1303.

35. Austevoll IM, Gjestad R, Brox JI, et al. The effectiveness of decompression alone compared with additional fusion for lumbar spinal stenosis with degenerative spondylolisthesis: a pragmatic comparative non-inferiority observational study from the Norwegian Registry for Spine Surgery. European Spine Journal. 2017;26(2):404–413.

36. Wiltse LL, Newman PH, Macnab I. Classification of spondylolisis and spondylolisthesis. Clin Orthop Relat Res. 1976;(117):23–29. NOT IN FILE.

37. Wiltse LL. Classification, terminology and measurements in spondylolisthesis. The Iowa orthopaedic journal. 1981;1:52.

38. Newman PH, Stone KH. The etiology of spondylolisthesis. Journal of Bone & Joint Surgery, British Volume. 1963;45(1):39–59. NOT IN FILE.

39. Fujiwara A, Tamai K, An HS, et al. The relationship between disc degeneration, facet joint osteoarthritis, and stability of the degenerative lumbar spine. JSpinal Disord. 2000;13(5):444–450. NOT IN FILE.

40. Chaput C, Padon D, Rush J, Lenehan E, Rahm M. The significance of increased fluid signal on magnetic resonance imaging in lumbar facets in relationship to degenerative spondylolisthesis. Spine. 2007;32(17):1883–1887. NOT IN FILE.

41. Frank DF, Miller JE. Hypoplasia of the lumbar vertebral body simulating spondylolisthesis. Radiology. 1979;133(1):59–60.

42. Niggemann P, Kuchta J, Grosskurth D, Beyer H, Hoeffer J, Delank K. Spondylolysis and isthmic spondylolisthesis: impact of vertebral hypoplasia on the use of the Meyerding classification. The British journal of radiology. 2012;85(1012):358–362.

43. Legaye J. Radiographic analysis of the listhesis associated with lumbar isthmic spondylolysis. Orthopaedics & Traumatology: Surgery & Research. 2018;104(5):569–573.

44. Mac-Thiong J-M, Labelle H, Parent S, Hresko MT, Deviren V, Weidenbaum M. Reliability and development of a new classification of lumbosacral spondylolisthesis. Scoliosis. 2008;3(1):1–9.

45. Wang YXJ, Kaplar Z, Deng M, Leung JC. Lumbar degenerative spondylolisthesis epidemiology: a systematic review with a focus on gender-specific and age-specific prevalence. Journal of orthopaedic translation. 2017;11:39–52.

46. Kalichman L, Kim DH, Li L, Guermazi A, Berkin V, Hunter DJ. Spondylolysis and spondylolisthesis: prevalence and association with low back pain in the adult community-based population. Spine. 2009;34(2):199.

47. Ishimoto Y, Cooper C, Ntani G, et al. Is radiographic lumbar spondylolisthesis associated with occupational exposures? Findings from a nested case control study within the Wakayama spine study. BMC Musculoskeletal Disorders. 2019;20(1):618.

48. Chen I-R, Wei T-S. Disc height and lumbar index as independent predictors of degenerative spondylolisthesis in middle-aged women with low back pain. Spine. 2009;34(13):1402–1409.

49. Shao Z, Rompe G, Schiltenwolf M. Radiographic changes in the lumbar intervertebral discs and lumbar vertebrae with age. Spine. 2002;27(3):263–268. NOT IN FILE.

50. Amonoo-Kuofi HS. Morphometric changes in the heights and anteroposterior diameters of the lumbar intervertebral discs with age. Journal of Anatomy. 1991;175:159. NOT IN FILE.

51. Twomey LT, Taylor JR. Age changes in lumbar vertebrae and intervertebral discs. Clinical Orthopaedics & Related Research. Nov 1987;(224):97–104. NOT IN FILE.

52. Videman T, Battié MC, Gibbons LE, Gill K. Aging changes in lumbar discs and vertebrae and their interaction: a 15-year follow-up study. The Spine Journal. 2014;14(3):469–478.

53. Bach K, Ford J, Foley R, et al. Morphometric analysis of lumbar intervertebral disc height: an imaging study. World neurosurgery. 2019;124:e106–e118.

54. Demir M, Emre A, Seringeç N, et al. Intervertebral disc heights and concavity index of the lumbar spine in young healthy adults. Anatomy. 2018;12(1):34–37.

55. Leone A, Cassar-Pullicino VN, Guglielmi G, Bonomo L. Degenerative lumbar intervertebral instability: what is it and how does imaging contribute? Skeletal radiology. 2009;38(6):529–533.

56. Lattig F, Fekete TF, Grob D, Kleinstuck FS, Jeszenszky D, Mannion AF. Lumbar facet joint effusion in MRI: a sign of instability in degenerative spondylolisthesis? European spine journal : official publication of the European Spine Society, the European Spinal Deformity Society, and the European Section of the Cervical Spine Research Society. Feb 2012;21(2):276–81. doi:10.1007/s00586-011-1993-1

57. Simmonds AM, Rampersaud YR, Dvorak MF, Dea N, Melnyk AD, Fisher CG. Defining the inherent stability of degenerative spondylolisthesis: a systematic review. Journal of Neurosurgery: Spine. 2015:1–12.

58. Hasegawa K, Kitahara K, Shimoda H, et al. Lumbar degenerative spondylolisthesis is not always unstable: clinicobiomechanical evidence. Spine. 2014;39(26):2127–2135.

59. Wang D, Yuan H, Liu A, et al. Analysis of the relationship between the facet fluid sign and lumbar spine motion of degenerative spondylolytic segment using Kinematic MRI. European journal of radiology. 2017;94:6–12.

60. Nizard RS, Wybier M, Laredo JD. Radiologic assessment of lumbar intervertebral instability and degenerative spondylolisthesis. The Radiologic clinics of North America. 2001;39(1):55–71.

61. Leone A, Guglielmi G, Cassar-Pullicino VN, Bonomo L. Lumbar intervertebral instability: a review. Radiology. Oct 2007;245(1):62–77. doi:10.1148/radiol.2451051359

62. Izzo R, Guarnieri G, Guglielmi G, Muto M. Biomechanics of the spine. Part I: spinal stability. Eur J Radiol. Jan 2013;82(1):118–26. doi:10.1016/j.ejrad.2012.07.024

63. Evans N, McCarthy M. Management of symptomatic degenerative low-grade lumbar spondylolisthesis. EFORT open reviews. 2018;3(12):620–631.

64. Wang M, Luo XJ, Ye YJ, Zhang Z. Does Concomitant Degenerative Spondylolisthesis Influence the Outcome of Decompression Alone in Degenerative Lumbar Spinal Stenosis? A Meta-Analysis of Comparative Studies. World neurosurgery. 2019;123:226–238.

65. Reitman CA, Cho CH, Bono CM, et al. Management of degenerative spondylolisthesis: development of appropriate use criteria. The Spine Journal. 2021;21(8):1256–1267.

66. Mannion A, Pittet V, Steiger F, Vader J-P, Becker H-J, Porchet F. Development of appropriateness criteria for the surgical treatment of symptomatic lumbar degenerative spondylolisthesis (LDS). European spine journal. 2014;23(9):1903–1917.

67. Even JL, Chen AF, Lee JY. Imaging characteristics of “dynamic” versus “static” spondylolisthesis: analysis using magnetic resonance imaging and flexion/extension films. The Spine Journal. 2014;14(9):1965–1969.

68. Inose H, Kato T, Onuma H, et al. Predictive Factors Affecting Surgical Outcomes in Patients with Degenerative Lumbar Spondylolisthesis. Spine. 2021;46(9):610–616.

69. Antani S, Cheng J, Long J, Long LR, Thoma GR. Medical validation and CBIR of spine x-ray images over the Internet. International Society for Optics and Photonics; 2006:60610J.

70. Korovessis P, Repantis T, Papazisis Z, Iliopoulos P. Effect of Sagittal Spinal Balance, Levels of Posterior Instrumentation, and Length of Follow-up on Low Back Pain in Patients Undergoing Posterior Decompression and Instrumented Fusion for Degenerative Lumbar Spine Disease: A Multifactorial Analysis. Spine. 2010;35(8):898.

71. Lord MJ, Small JM, Dinsay JM, Watkins RG. Lumbar lordosis. Effects of sitting and standing. Spine. 1997;22(21):2571–2574. NOT IN FILE.

72. Been E, Barash A, Pessah H, Peleg S. A new look at the geometry of the lumbar spine. Spine. 2010;35(20):E1014–E1017.

73. Crawford NR, Peles JD, Dickman CA. The spinal lax zone and neutral zone: measurement techniques and parameter comparisons. J Spinal Disord. Oct 1998;11(5):416–29.

74. Panjabi MM. The stabilizing system of the spine. Part II. Neutral zone and instability hypothesis. J Spinal Disord. Dec 1992;5(4):390–6; discussion 397. NOT IN FILE. doi:10.1097/00002517-199212000-00002

75. Black DM, Cummings SR, Stone K, Hudes E, Palermo L, Steiger P. A new approach to defining normal vertebral dimensions. J Bone Min Res. 1991;6:883–892. NOT IN FILE.

76. Rea J, Steiger P, Blake G, Potts E, Smith I, Fogelman I. Morphometric X-ray absorptiometry: reference data for vertebral dimensions. Journal of Bone and Mineral Research. 1998;13(3):464–474.

77. O’Neill T, Varlow J, Felsenberg D, et al. Variation in vertebral height ratios in population studies. J Bone Miner Res. 1994;12:1895–1907. NOT IN FILE.

## References

1. Hurxthal LM. Measurement of anterior vertebral compressions and biconcave vertebrae. Am J Roentgenol Radium Ther Nucl Med. Jul 1968;103(3):635–44.

2. Diacinti D, Guglielmi G. Vertebral morphometry. Radiologic clinics of North America. May 2010;48(3):561–75.

3. Frobin W, Brinckmann P, Biggemann M, Tillotson M, Burton K. Precision measurement of disc height, vertebral height and sagittal plane displacement from lateral radiographic views of the lumbar spine. Clin Biomech (Bristol, Avon). 1997;12 Suppl 1:S1–S63.

4. Keynan O, Fisher CG, Vaccaro A, Fehlings MG, Oner FC, Dietz J, et al. Radiographic measurement parameters in thoracolumbar fractures: a systematic review and consensus statement of the spine trauma study group. Spine (Phila Pa 1976). 10.1097/01.brs.0000201261.94907.0d doi;00007632-200603010-00025 pii Mar 1 2006;31(5):E156–65.

5. Quint DJ, Tuite GF, Stern JD, Doran SE, Papadopoulos SM, McGillicuddy JE, et al. Computer-assisted measurement of lumbar spine radiographs. Academic radiology. 1997;4(11):742–52.

6. Brinckmann P, Frobin W, Biggemann M, Hilweg D, Seidel S, Burton K, et al. Quantification of overload injuries to thoracolumbar vertebrae and discs in persons exposed to heavy physical exertions or vibration at the work-place: The shape of vertebrae and intervertebral discs—study of a young, healthy population and a middle-aged control group. Clinical Biomechanics. 1994;9:S3–S83.

## References

1. Mummaneni PV, Bisson EF, Kerezoudis P, et al. Minimally invasive versus open fusion for Grade I degenerative lumbar spondylolisthesis: analysis of the Quality Outcomes Database. Neurosurgical focus. 2017;43(2):E11.

2. Staub BN, Holman PJ, Reitman CA, Hipp JA. Sagittal plane lumbar intervertebral motion during seated flexion-extension in 658 asymptomatic, non-degenerated levels. J Neurosurgery Spine. 2015 2015;23(6):731–738. doi:10.3171/2015.3.SPINE14898

3. Kellgren J, Lawrence J. Radiological assessment of osteo-arthrosis. Annals of the Rheumatic Diseases. 1957;16(4):494.

4. Kellgren J, Lawrence J. Atlas of standard radiographs: the epidemiology of chronic rheumatism. Vol. 2. Oxford: Blackwell; 1963.

## References

1. Fedorov A, Beichel R, Kalpathy-Cramer J, et al. 3D Slicer as an image computing platform for the Quantitative Imaging Network. Magnetic resonance imaging. 2012;30(9):1323–1341. doi:https://doi.org/10.1016/j.mri.2012.05.001

2. Shackleford JA, Shusharina N, Verberg J, et al. Plastimatch 1.6-current capabilities and future directions. 2012:

3. Andersson G, Schultz A, Nathan A, Irstam L. Roentgenographic measurement of lumbar intervertebral disc height. Spine. 1981;6(2):154–158.

4. Saraste H, Brostrom LA, Aparisi T, Axdorph G. Radiographic measurement of the lumbar spine. A clinical and experimental study in man. Spine. 1985;10(3):236–241. NOT IN FILE.

5. Frobin W, Brinckmann P, Biggemann M, Tillotson M, Burton K. Precision measurement of disc height, vertebral height and sagittal plane displacement from lateral radiographic views of the lumbar spine. Clinical Biomechanics. 1997;12:S1–S63.

